# Combining evidence from human genetic and functional screens to identify pathways altering obesity and fat distribution

**DOI:** 10.1101/2024.09.19.24313913

**Authors:** Nikolas A. Baya, Ilknur Sur Erdem, Samvida S. Venkatesh, Saskia Reibe, Philip D. Charles, Elena Navarro-Guerrero, Barney Hill, Frederik Heymann Lassen, Melina Claussnitzer, Duncan S. Palmer, Cecilia M. Lindgren

## Abstract

Overall adiposity and body fat distribution are heritable traits associated with altered risk of cardiometabolic disease and mortality. Performing rare variant (minor allele frequency<1%) association testing using exome-sequencing data from 402,375 participants in the UK Biobank (UKB) for nine overall and tissue-specific fat distribution traits, we identified 19 genes where putatively damaging rare variation associated with at least one trait (Bonferroni-adjusted *P*<1.58×10^-7^) and 50 additional genes at FDR≤1% (*P*≤4.37×10^-5^). These 69 genes exhibited significantly higher (one-sided *t*-test *P*=3.58×10^-18^) common variant prioritisation scores than genes not significantly enriched for rare putatively damaging variation, with evidence of monotonic allelic series (dose-response relationships) among ultra-rare variants (minor allele count≤10) in 22 genes. Combining rare and common variation evidence, allelic series and longitudinal analysis, we selected 14 genes for CRISPR knockdown in human white adipose tissue cell lines. In three previously uncharacterised target genes, knockdown increased (two-sided *t*-test *P*<0.05) lipid accumulation, a cellular phenotype relevant for fat mass traits, compared to Cas9-empty negative controls: *COL5A3* (fold change [FC]=1.72, *P*=0.0028), *EXOC7* (FC=1.35, *P*=0.0096), and *TRIP10* (FC=1.39, *P*=0.0157); furthermore, knockdown of *PPARG* (FC=0.25, *P*=5.52×10^-7^) and *SLTM* (FC=0.51, *P*=1.91×10^-4^) resulted in reduced lipid accumulation. Integrating across population-based genetic and *in vitro* functional evidence, we highlight therapeutic avenues for altering obesity and body fat distribution by modulating lipid accumulation.

## Introduction

One in four adults globally are either overweight or obese^1,2^. While higher overall adiposity increases morbidity and mortality,^1,3^ disease risk is also informed by the location and distribution of excess fat within particular depots^4,5^. Abnormal distribution of fat is often attributed to lipodystrophy syndromes, which can cause generalised or selective fat mass loss and depot-specific fat growth^6^. Independent of overall body mass index (BMI), individuals with higher central adiposity have increased risk of cardiometabolic diseases, including type 2 diabetes (T2D) and stroke^7,8^; in contrast, individuals with higher hip circumference, an indicator for gluteal adiposity, have lower risk of such outcomes. For example, a standard deviation increase in hip circumference has been shown to reduce risk of T2D by ∼40%^9^ or myocardial infarction by ∼10%^10^. Previous studies indicate that fat distribution, as assessed by waist-to-hip ratio (WHR), has a strong heritable component independent of BMI, with narrow sense heritability of up to 56% in women and 32% in men^8,11^.

BMI-associated genes are enriched in tissues of the central nervous system, notably the hypothalamus which is involved in appetite regulation^12^. Indeed, blockbuster GLP1-receptor agonists that are prescribed for weight loss^13,14^ act primarily through the dorsomedial hypothalamus to control food intake^15^. In contrast, genome-wide association studies (GWASs) for WHR adjusted for BMI (WHRadjBMI) indicate enrichment of genes associated with fat distribution in adipose tissue^16^. However, there are currently few therapeutic avenues to modify both obesity and fat distribution^17^.

Understanding the genetic aetiology of body fat and fat distribution may drive new therapies for obesity. Mendelian genetic studies have identified rare variants in genes such as *PPARG*, a master regulator of adipocyte differentiation, and *INSR*, the insulin receptor, associated with lipodystrophies and extreme forms of central body fat distribution^18,19^. In the general population, rare protein-coding variants with large effects can point to genetic and molecular mechanisms underpinning fat distribution. For example, previous work has demonstrated that rare loss-of-function alleles in *GPR75* are protective against obesity^20^, and protein-truncating variants in *INHBE* are associated with favourable fat distribution^21^.

Indeed, while there have been several common and rare-variant association studies of obesity^16,20–24^, mechanistic studies of the pathways by which the identified genes affect adipogenesis have typically been limited to characterising just one or two targets^25–33^. The difficulty of manipulating human adipocytes in high-throughput experimental assays has prevented genome-wide functional studies of the genetic drivers of adipogenesis.

Here, we integrated exome-sequencing (ES) and common variant GWASs for nine obesity-related and fat distribution traits to nominate genes associated with overall obesity or central adiposity in up to 402,375 participants in UKB. We then designed an *in vitro* functional assay for lipid accumulation in human white adipocytes to biologically validate a considerable selection of the identified genes using CRISPR-Cas9 knockdown experiments. Taken together, we demonstrate that converging multi-modal evidence from GWAS, ES, and *in vitro* knockdown studies in a relevant context can generate putative therapeutic targets for obesity or lipodystrophy.

## Results

### Discovery of genes associated with obesity and fat distribution through exome-wide association tests

We performed gene-level discovery testing across 1,827,504 rare variants (minor allele frequency [MAF] <1%) in 18,788 genes for association with three obesity or fat distribution related traits, i.e. overall obesity (BMI and body fat percentage) and central fat distribution (WHRadjBMI) in up to 402,375 participants of European ancestry in the UKB (Supp. Tables 1 and 2). We additionally assessed the effects of these genes on tissue-specific fat component phenotypes derived from dual energy X-ray absorptiometry (DXA) and magnetic resonance imaging (MRI) scans (such as android tissue fat percentage, abdominal fat ratio, and visceral adipose tissue volume), in up to 39,671 participants of European ancestry in the UKB (Supp. Table 2). We defined damaging rare variants as those annotated to be high-confidence predicted loss-of-function (pLoF) or damaging missense using a combination of existing effect prediction tools (LOFTEE^34^, CADD^35^, REVEL^36^, and SpliceAI^37^; Methods). The reported gene-level effect sizes were estimated by burden testing, while *P*-values were estimated using the optimal sequence kernel association test (SKAT-O) for improved power (Methods). Finally, to account for potential confounding due to nearby common variants, we conditioned gene-level associations on fine-mapped GWAS loci on the same chromosome (Methods).

**Table 1.** Exome-wide significant gene-level associations. Gene-level exome-wide significance threshold: SKAT-O *P*<1.58×10^-7^. Genomic location indicates the chromosome and base pair coordinates for the start of the gene in Genome Reference Consortium Human Build 38. Variants in the mask are those which are included in the SKAT-O test for a given gene. They include all variants in the gene with MAF<1% and consequence annotation matching the specific mask. Total MAF is the sum of MAF for all variants in the mask. In parentheses, MAC is the total minor allele count for the gene among individuals included in the association test. The effect size of a gene is estimated using burden testing and is in units of the phenotype’s standard deviation. The 95% confidence interval is defined by an interval centred on the effect size point estimate, +/− 1.96 standard errors. The unweighted effect size is a version of the burden effect size that does not weight variants by Beta(MAF; 1, 25), the default weighting used by SAIGE. Using the unweighted effect sizes puts the gene-level effect on the same scale as variant-level effects. See Supp. Note 1 for calculation of unweighted effect size. WHRadjBMI, waist-to-hip ratio adjusted for BMI; BMI, body mass index; pLoF, predicted loss-of-function.

| Gene<br>(genomic<br>location) | Phenotype | Consequence | Variants in<br>mask | Cumulative<br>MAF (MAC) | SKAT-O<br>P-value | Burden<br>P-value | Effect size<br>[95% CI] | Effect size,<br>unweighted |
| --- | --- | --- | --- | --- | --- | --- | --- | --- |
| *ANKRD12*<br>(18:9136228) | WHRadjBMI | pLoF | 127 | 0.00038<br>(308) | $1.71 \times 10^{-9}$ | $6.67 \times 10^{-8}$ | 0.010<br>[0.006, 0.013] | 0.238 |
| *APBA1*<br>(9:69427532) | BMI | pLoF | 59 | 0.00016<br>(128) | $6.25 \times 10^{-8}$ | $5.18 \times 10^{-8}$ | 0.019<br>[0.012, 0.026] | 0.475 |
| *BLTP1*<br>(4:122152331) | BMI | pLoF | 282 | 0.00075<br>(600) | $2.15 \times 10^{-10}$ | $9.19 \times 10^{-11}$ | 0.011<br>[0.007, 0.014] | 0.258 |
| *COL5A3*<br>(19:9959561) | WHRadjBMI | pLoF plus damaging<br>missense | 362 | 0.00388<br>(3,111) | $5.23 \times 10^{-10}$ | $2.98 \times 10^{-9}$ | 0.003<br>[0.002, 0.004] | 0.078 |
| *DIDO1*<br>(20:62877738) | BMI | pLoF | 22 | 0.00003<br>(25) | $1.14 \times 10^{-7}$ | $1.14 \times 10^{-7}$ | 0.042<br>[0.026, 0.057] | 1.050 |
| *GPR151*<br>(5:146513144) | Body fat % | pLoF | 37 | 0.01039<br>(8,209) | $6.60 \times 10^{-8}$ | $2.51 \times 10^{-7}$ | -0.002<br>[-0.003, -0.001] | -0.043 |
| | BMI | pLoF | 37 | 0.01036<br>(8,313) | $1.35 \times 10^{-7}$ | $2.02 \times 10^{-7}$ | -0.003<br>[-0.004, -0.002] | -0.058 |
| *INHBE*<br>(12:57452323) | WHRadjBMI | pLoF | 23 | 0.00119<br>(954) | $3.27 \times 10^{-9}$ | $1.37 \times 10^{-7}$ | -0.005<br>[-0.007, -0.003] | -0.123 |
| *INSR*<br>(19:7112255) | WHRadjBMI | pLoF | 55 | 0.00019<br>(151) | $6.38 \times 10^{-8}$ | $4.20 \times 10^{-7}$ | -0.012<br>[-0.017, -0.007] | -0.299 |
| *KEAP1*<br>(19:10486125) | WHRadjBMI | pLoF plus damaging<br>missense | 88 | 0.00034<br>(273) | $1.76 \times 10^{-11}$ | $2.52 \times 10^{-12}$ | 0.012<br>[0.009, 0.016] | 0.305 |
| *MC4R*<br>(18:60371062) | BMI | pLoF | 23 | 0.00023<br>(187) | $9.30 \times 10^{-13}$ | $1.87 \times 10^{-13}$ | 0.021<br>[0.016, 0.027] | 0.532 |
| *PDE3B*<br>(11:14643804) | WHRadjBMI | pLoF | 68 | 0.00132<br>(1,057) | $5.91 \times 10^{-10}$ | $7.09 \times 10^{-10}$ | -0.007<br>[-0.009, -0.005] | -0.166 |
| | Body fat % | pLoF | 66 | 0.00132<br>(1,046) | $1.34 \times 10^{-7}$ | $2.09 \times 10^{-7}$ | 0.005<br>[0.003, 0.007] | 0.102 |
| *PKD1*<br>(16:2088708) | WHRadjBMI | pLoF | 97 | 0.00022<br>(176) | $7.68 \times 10^{-11}$ | $8.50 \times 10^{-7}$ | 0.012<br>[0.007, 0.017] | 0.311 |
| *PLD1*<br>(3:171600404) | WHRadjBMI | pLoF plus damaging<br>missense | 240 | 0.01163<br>(9,324) | $4.69 \times 10^{-10}$ | $2.88 \times 10^{-9}$ | 0.002<br>[0.001, 0.003] | 0.044 |
| *PLIN1*<br>(15:89664367) | WHRadjBMI | pLoF | 48 | 0.00104<br>(836) | $8.94 \times 10^{-23}$ | $8.98 \times 10^{-21}$ | -0.009<br>[-0.011, -0.007] | -0.233 |
| *PLIN4*<br>(19:4502192) | WHRadjBMI | pLoF | 121 | 0.00444<br>(3,560) | $1.00 \times 10^{-13}$ | $3.25 \times 10^{-14}$ | 0.004<br>[0.003, 0.005] | 0.091 |
| | Body fat % | pLoF | 120 | 0.00444<br>(3,506) | $1.23 \times 10^{-10}$ | $2.11 \times 10^{-10}$ | -0.003<br>[-0.004, -0.002] | -0.080 |
| *PPARG*<br>(3:12287368) | Body fat % | pLoF plus damaging<br>missense | 66 | 0.00021<br>(168) | $2.90 \times 10^{-8}$ | $4.34 \times 10^{-7}$ | -0.012<br>[-0.016, -0.007] | -0.292 |
| *PTPRG*<br>(3:61561569) | BMI | pLoF | 120 | 0.00024<br>(194) | $1.37 \times 10^{-8}$ | $8.31 \times 10^{-8}$ | 0.015<br>[0.01, 0.021] | 0.377 |
| | Body fat % | pLoF | 120 | 0.00024<br>(192) | $1.35 \times 10^{-7}$ | $8.25 \times 10^{-7}$ | 0.011<br>[0.006, 0.015] | 0.267 |
| *RIF1*<br>(2:151409883) | Body fat % | pLoF | 66 | 0.00011<br>(89) | $1.41 \times 10^{-7}$ | $1.41 \times 10^{-7}$ | 0.017<br>[0.01, 0.023] | 0.416 |
| *SLC12A5*<br>(20:46021690) | BMI | pLoF | 13 | 0.00002<br>(13) | $1.21 \times 10^{-7}$ | $1.21 \times 10^{-7}$ | 0.058<br>[0.036, 0.079] | 1.446 |

We identified 19 unique genes carrying rare damaging variation associated with BMI, body fat percentage, or WHRadjBMI at exome-wide significance (SKAT-O *P*<1.58×10^-7^, Bonferroni adjustment for 315,996 unique tests) (Table 1 and Figure 1a). Four genes associated with body fat percentage are also exome-wide significant for BMI with the same direction of effect (*PTPRG* and *GPR151*) or WHRadjBMI with opposing directions of effect (*PDE3B* and *PLIN4*) (Figure 2, Supp. Figure 3). *PLIN4* is also negatively associated with gynoid fat percentage (SKAT-O *P*=2.16×10^-6^), suggesting that the association of *PLIN4* with lower body fat percentage may be driven by reduced gynoid fat deposits, which in turn results in a positive association with WHRadjBMI (an anthropometric proxy for the ratio of android to gynoid fat accumulation). We see a qualitatively similar effect of a burden of rare variation in *PDE3B*, which is associated with higher overall obesity as measured by body fat percentage, but reduced WHRadjBMI (exome-wide significant), android to gynoid ratio, abdominal fat ratio, and visceral adipose tissue volume (all *P*<0.05), indicating that the increased body fat percentage in these individuals may be driven by higher subcutaneous rather than visceral fat accumulation (Figure 2, Supp. Figure 3).

**Figure 1.**
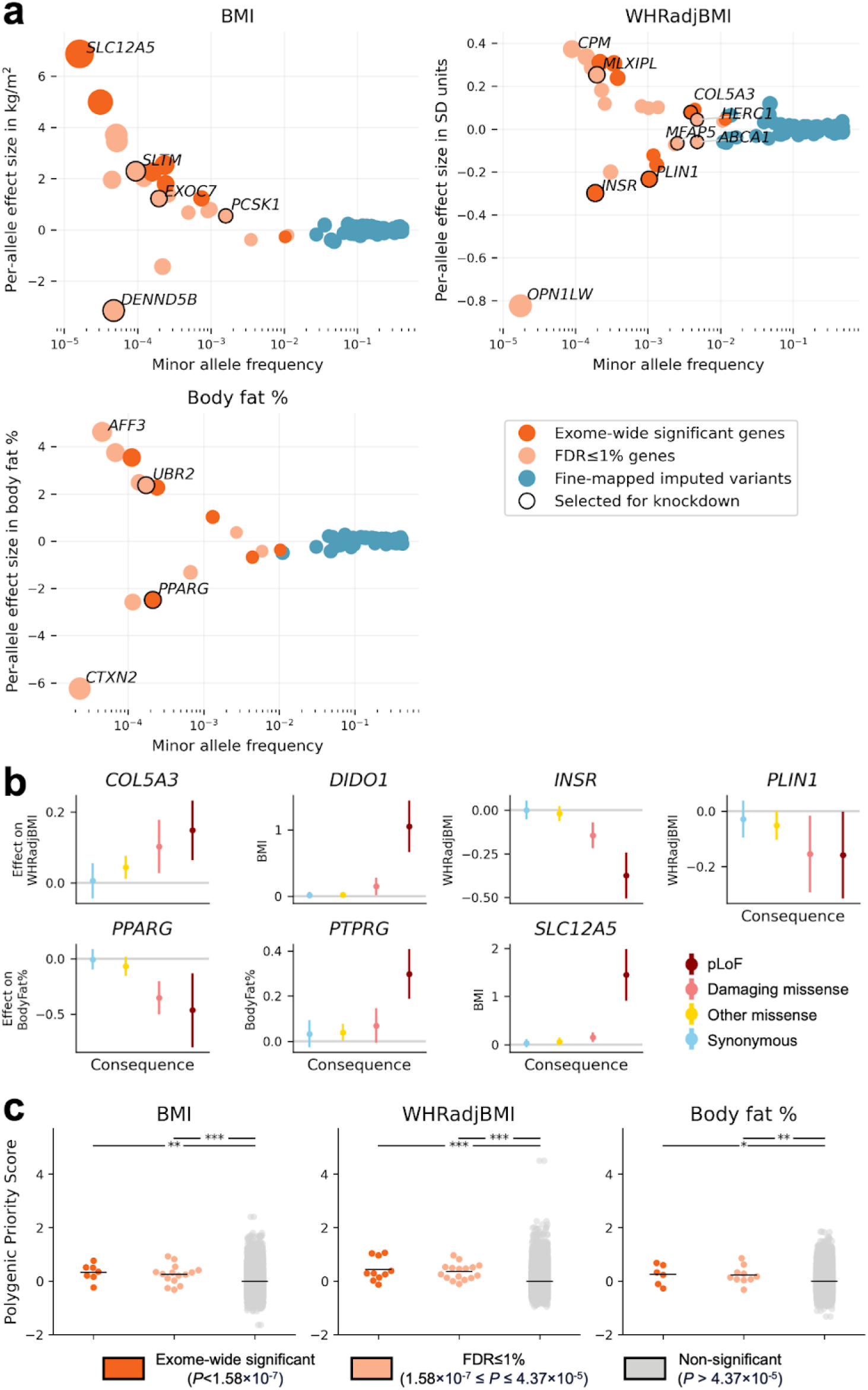
Consequences of rare missense variation on obesity or fat distribution related traits in UKB. **a**, Gene– and variant-level effects as a function of aggregated minor allele frequency. Only gene-level results which are exome-wide or FDR≤1% significant are shown. Only fine-mapped common (MAF>1%) variants with posterior inclusion probability ≥0.9 are shown. Genes selected for functional characterisation by knockdown are circled in black. Effect sizes for BMI and body fat percentage are converted to the kg/m^2^ and fat percentage scale, respectively, by multiplying the effect size in standard deviation (SD) units by the empirical SD of each trait (4.75 kg/m^2^ for BMI, 8.51% for body fat percentage). Genes selected for knockdown are outlined in black and labeled. Genes with highest and lowest effect size for each trait are also labeled. Point sizes are scaled by effect size. **b,** Allelic series for exome-wide significant genes with monotonic relationships between effect size and consequence severity in ultra-rare (MAC≤10) variant burden tests. Confidence intervals for effect size defined as +/− 1.96 standard errors. **c,** Enrichment of Polygenic Priority Scores (PoPS)^38^ among exome-wide and FDR≤1% significant genes compared to other genes. Significance of one-sided *t*-test with alternate hypothesis that a significant gene category has higher average PoPS than the non-significant gene category: \*\*\**P*<0.001, \*\**P*<0.01, \**P*<0.05. Results of *t*-test for BMI PoPS: exome-wide significant gene PoPS vs. non-significant gene PoPS *t*-statistic (*P*)=2.79 (2.65×10^-3^), FDR≤1% vs. non-significant=3.17 (7.64×10^-4^). Results of *t*-test for WHRadjBMI PoPS: exome-wide significant vs. non-significant=4.96 (3.60×10^-7^), FDR≤1% vs. non-significant=5.19 (1.06×10^-7^). Results of *t*-test for body fat percentage PoPS: exome-wide significant vs. non-significant=2.04 (2.07×10^-2^), FDR≤1% vs. non-significant=2.46 (7.02×10^-3^). The mean PoPS for each gene category are indicated by black bars. For BMI: Exome-wide significant genes PoPS mean [95% CI from +/− 1.96 SEM] (number of genes)=0.324 [0.086, 0.562] (7), FDR≤1%=0.250 [0.070, 0.430] (15), non-significant=-0.009 [-0.012, –0.006] (34613). For WHRadjBMI: Exome-wide significant=0.430 [0.161, 0.698] (10), FDR≤1%=0.353 [0.207, 0.500] (16), non-significant=-0.014 [-0.017, –0.011] (34609). For body fat percentage: Exome-wide significant=0.242 [-0.060, 0.544] (6), FDR≤1%=0.226 [0.023, 0.429] (10), non-significant=-0.006 [-0.009, –0.003] (34619).

**Figure 2.**
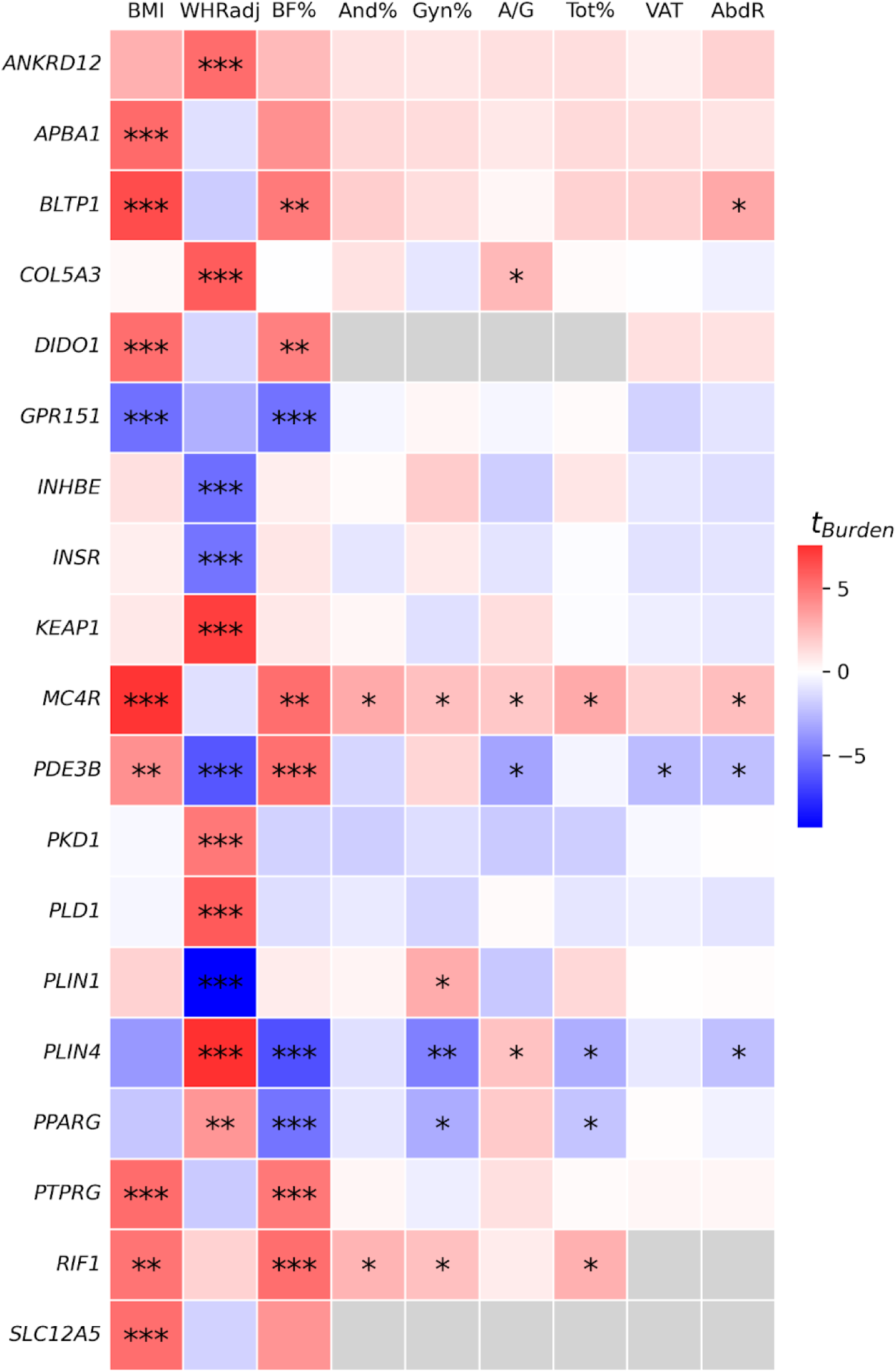
Gene burden effects across all nine obesity and fat distribution traits for 19 genes with exome-wide significant burden associations. Heatmap is coloured by the burden test *t*-statistic. Only the result of the consequence mask with the lowest SKAT-O *P* is shown for each trait-gene pair. Significance of SKAT-O test: ***Exome-wide significant (*P*<1.58×10^-7^), **FDR≤1% significant (*P*≤4.37×10^-5^), *Nominal significance (*P*<0.05). Grey squares indicate that the gene-level test could not be performed due to insufficient allele counts. WHRadj, waist-to-hip ratio adjusted for BMI. BF%, body fat percentage. And%, android fat percentage. Gyn%, gynoid fat percentage. A/G, android-gynoid fat percentage ratio. Tot%, total fat percentage. VAT, visceral adipose tissue. AbdR, abdominal fat ratio.

While the 19 exome-wide significant genes were not significantly associated (all *P*>4.38×10^-4^, Bonferroni adjustment for 19 unique genes x 6 phenotypes) with any other tissue-specific fat components, we observed that nominally significant associations (*P*<0.05) generally shared the expected direction of effect on the primary and supplemental phenotypes (Figure 2, Supp. Figure 3). Genes that are positively associated with BMI (*MC4R*) and body fat percentage (*RIF1*) are also positively associated with android fat percentage, gynoid fat percentage, android/gynoid ratio, and total tissue fat percentage; and *PPARG* is negatively associated with body fat percentage as well as gynoid and total tissue fat percentage (Figure 2, Supp. Figure 3). Similarly, *COL5A3* is associated with increased WHRadjBMI and higher android to gynoid ratio, while *PLIN1* is associated with lower WHRadjBMI and higher gynoid fat percentage as expected.

Finally, we found evidence of monotonic allelic series, that is, increasingly large effects of increasingly damaging variants in a dose-response relationship, among ultra-rare variants (minor allele count (MAC)≤10) in *COL5A3, DIDO1, INSR, PLIN1, PTPRG, PPARG* and *SLC12A5* (Figure 1b).

### Convergence of rare and common variant evidence for genes associated with obesity and fat distribution

Genes associated with obesity and fat distribution in rare variant gene-level tests overlapped with those highlighted by common variant associations, demonstrated by a significant enrichment of the Polygenic Priority Score (PoPS)^38^ (one-sided *t*-test *P*<0.05, Figure 1c) among genes harbouring exome-wide significant damaging rare variant association signals. Moreover, 50 rare-variant gene-level associations which do not reach exome-wide significance but pass a less stringent significance threshold (SKAT-O *P*≤4.37×10^-5^, using the Benjamini-Hochberg procedure^39^ for FDR≤1% on minimum SKAT-O *P*-value across traits and two variant masks; Methods) also have significantly higher PoPS compared to non-significant genes (one-sided *t*-test *P*<0.01, Figure 1c), indicating value in examining these additional genes as potential therapeutic targets for obesity and lipodystrophy (Supp. Table 3). Seven additional gene-level associations also passed the FDR≤1% significance threshold, but were supported by minor allele counts less than ten (Supp. Note 2). Combining PoPS across all gene-trait pairs (for traits with PoPS available: BMI, WHRadjBMI, body fat percentage) which are exome-wide or FDR≤1% significant and comparing to PoPS of non-significant gene-trait pairs, we observe strong enrichment of scores (one-sided *t*-test: *t*=8.47, *P*=1.26×10^-17^, mean [95% CI using +/− 1.96྾SEM] PoPS of 64 exome-wide or FDR≤1% significant gene-trait pairs=0.307 [0.223, 0.392], mean of 103,841 non-significant gene-trait pairs=-0.010 [-0.012, –0.008]), demonstrating again the convergence of rare and common variant evidence when FDR≤1% significant genes are included.

Of the 31 genes known to cause severe monogenic obesity or lipodystrophy^6,24^, three (*PPARG*, *PLIN1*, and *MC4R*) are associated with an obesity or fat distribution trait at the exome-wide level (*P<*1.58×10^-7^) in our rare-variant gene-level association testing, representing a 125-fold enrichment over random chance (Fisher’s exact test *P*=3.87×10^-6^). Of the 69 FDR-significant genes, 23 have previously been reported to have rare variant gene-level associations with traits related to obesity or fat distribution in UKB participants, across a variety of variant annotation and gene-level association testing methods (Supp. Table 8)^20–22,40^. We looked up the effects of pLoF variant burden in 68/69 FDR-significant genes with MAC≥10 in the multi-ancestry meta-analysed All of Us tables for available phenotypes: BMI and WHRadjBMI^41^. Accounting for the multiple testing burden of 68 genes and two phenotypes (Bonferroni adjustment, *P*<3.68×10^-4^), we replicated, with matching effect directions, the associations of *SLTM*, *MC4R*, and *KIF1B* with BMI, and *PKD1* and *PLIN4* with WHRadjBMI (Supp. Table 4). We also found that *HECTD4* is positively associated with body fat percentage in UKB and BMI in All of Us; and an additional 18 of our FDR-significant genes are nominally associated (*P*<0.05) with an obesity-related trait in All of Us (Supp. Table 4).

Lastly, to evaluate the phenome-wide effects of genes related to obesity and fat distribution, we scanned 4,529 phenotypes in Genebass^42^ summary statistics for associations with the 69 FDR-significant genes. We found 549 significant associations (Genebass SKAT-O *P*≤9.98×10^-6^, FDR≤1%) across 211 traits for 41/69 genes (Supp. Figure 5, Supp. Table 5). The most significant Genebass associations were among blood biochemistry lipid phenotypes (Supp. Figure 5a, Supp. Table 5). We also observed shared associations across FDR-significant genes for phenotypes significantly genetically correlated to BMI and body fat percentage, such as fat mass and distribution traits. For example, eight FDR-significant genes (*BLTP1, DIDO1, GPR151, MC4R, PDE3B, PLIN4, RIF1, UBR2*) are associated with arm fat mass, leg fat mass, arm fat percentage, whole body fat mass, and hip circumference (all traits have common variant LD Score regression^43^ *r*_g_>0.80, *P*≤1×10^-308^ with BMI and body fat percentage, Supp. Table 5)^44^. We see some evidence for pleiotropy, as defined by associations with phenotypes genetically uncorrelated with BMI or body fat percentage, among results with common variant genetic correlations available: *PDE3B* and *STAB1* are associated with platelet distribution width (BMI *r*_g_=0.025, *P*=0.274; body fat percentage *r*_g_=9.7×10^-4^, *P*=0.958) and *ANKRD12* is associated with neutrophil percentage (BMI *r*_g_=-0.029, *P*=0.117; body fat percentage *r*_g_=-8.8×10^-3^, *P*=0.639) (Supp. Table 5).

### Sex– and age-specific effects of genes associated with obesity and fat distribution

We performed all association tests in sex-specific strata, identifying three genes (*INSR, PDE3B, PLIN4*) with significant differences (sex-difference two-sided heterogeneity test of effect sizes *P*<2.67×10^-6^, Bonferroni adjusted for 18,737 genes tested for sex-differential effects; Methods) in burden effect sizes. We observed opposing sex-specific effects for *INSR* on WHRadjBMI (female burden beta (SE)=-0.00458 (9.44×10^-4^), male beta=0.00195 (0.00100), sex-difference *P*=1.66×10^-6^). We found that female-specific effects may be driving the reported sex-combined associations between WHRadjBMI and *PDE3B* (female beta=-0.0169 (0.00169), male beta=-0.00288 (0.00179), sex-difference *P*=8.02×10^-9^) and *PLIN4* (female beta=0.00822 (9.15×10^-4^), male beta=0.00108 (9.86×10^-4^), sex-difference *P*=8.19×10^-8^) (Supp. Figure 6a, Supp. Table 6). Within the sex-specific analysis, we find two genes with female-specific associations (*P*<2.67×10^-6^, Bonferroni adjusted for 18,737 genes tested for sex-specific effects) of pLoF variation on obesity and fat distribution (*CASQ1* with body fat percentage, female SKAT-O *P*=1.12×10^-6^; *PLXND1* with WHRadjBMI, female *P*=9.69×10^-10^), and two genes with male-specific associations, both for the ratio of android to gynoid tissue fat percentage (*ZNF841* male *P*=1.04×10^-6^; *TINF2* male *P*=4.84×10^-7^) (Supp. Figure 6b, Supp. Table 6), which do not reach significance at the FDR≤1% threshold (*P*≤4.37×10^-5^) in sex-combined analyses.

We leveraged the age at diagnosis of obesity in longitudinal health records linked to UKB participants to identify that a burden of rare missense variation (MAF<1%) in five genes was associated (*P*<2.42×10^-4^, Bonferroni adjusted for 207 gene-consequence mask pairs) with elevated lifetime risk of obesity (Supp. Figure 7, Supp. Table 7). Individuals carrying rare missense variants in *MC4R* (Cox proportional hazard ratio [HR] (SE)=1.46 (0.0843), *P*=7.75×10^-6^), men with pLoF variants in *SLTM* (HR=5.37 (0.354), *P*=2.05×10^-6^) and damaging missense variants in *PCSK1* (HR=1.86 (0.165), *P*=1.63×10^-4^), and women with pLoF variants in *DIDO1* (HR=11.2 (0.578), *P*=2.85×10^-5^) and *SLC12A5* (HR=14.8 (0.708), *P*=1.45×10^-4^) were at risk of earlier age at onset of obesity (Supp. Figure 7, Supp. Table 7).

### Nominating target genes for *in vitro* functional characterisation through CRISPR knockdown

As adipose tissue is enriched for the expression of fat distribution genes identified from common variant GWASs^16^, we assessed the mRNA counts of overall and tissue-specific fat distribution-associated genes from our exome-wide gene-level association testing in differentiated human white adipose tissue derived pre-adipocytes (hWAT) *in vitro* (Figure 3a). Genes with FDR-significant results in gene-level tests had higher expression in hWAT 8 and 24 days after differentiation compared to non-significant genes (one-sided *t*-test *P*=1.55×10^-43^, comparing mRNA normalised counts). From the 56 of 69 FDR-significant genes sufficiently expressed in differentiated hWAT (mean normalised mRNA count >10 at 8 and 24 days after differentiation), we selected for functional characterisation 14 genes (*ABCA1, COL5A3, DENND5B, EXOC7, HERC1, INSR, MFAP5, MLXIPL, PCSK1, PLIN1, PPARG, SLTM, TRIP10*, *UBR2*; Figure 3b) which: (1) demonstrated monotonic allelic series indicating a dose-response relationship with gene dosage (*ABCA1*, *COL5A3, DENND5B, INSR, PCSK1, PLIN1, PPARG*) (Supp. Figure 5), (2) associated with obesity age-of-onset (*SLTM, PCSK1*) (Supp. Figure 7, Supp. Table 7), or (3) are implicated in lipid or glucose metabolism pathways by mouse whole-body or adipocyte-specific gene perturbation (Supp. Table 8).

**Figure 3.**
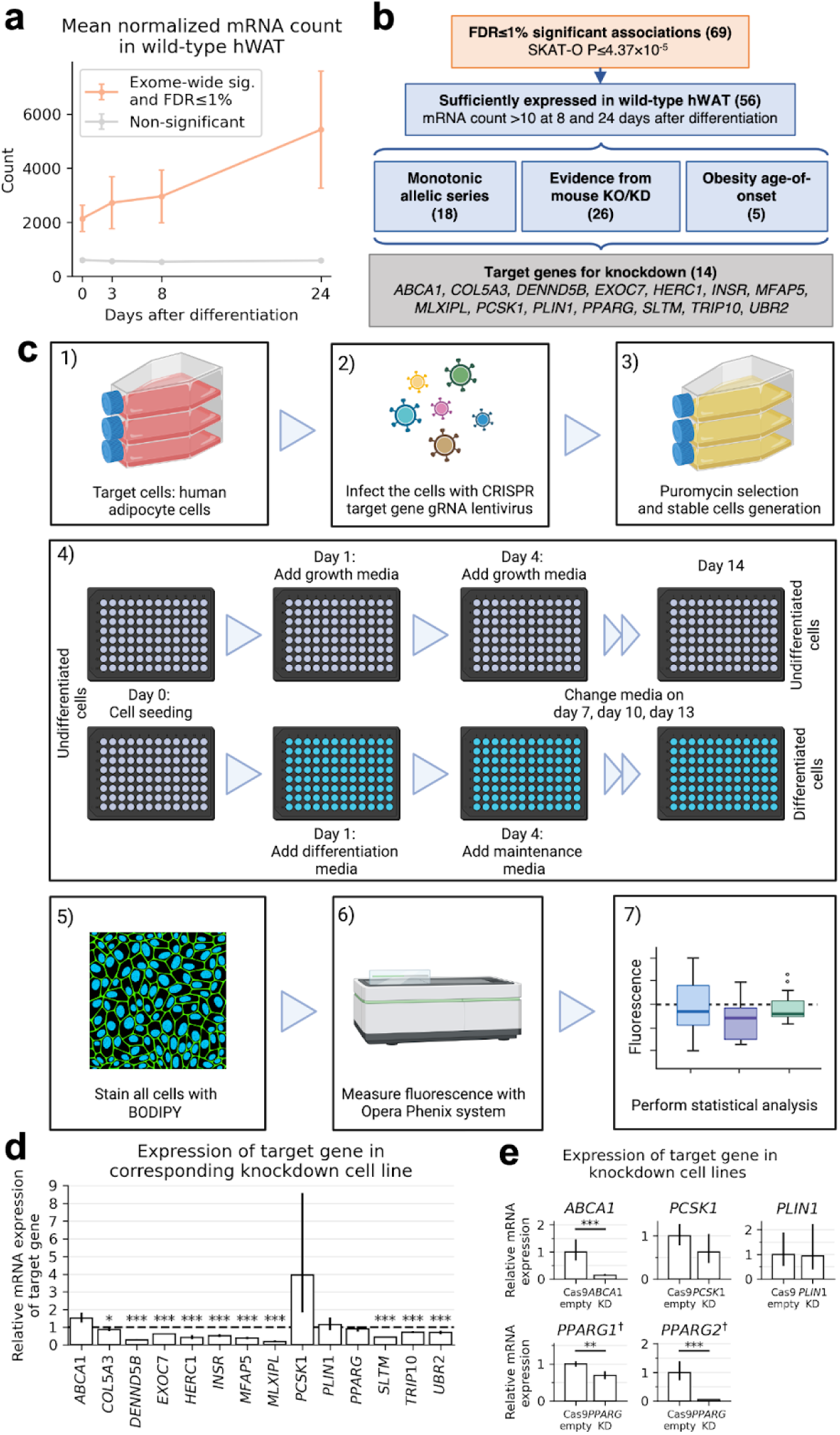
CRISPR knockdown strategy. **a**, Mean normalised mRNA counts across four time points (days after differentiation: 0, 3, 8, 24) in human white adipocyte tissue for 69 genes passing FDR≤1% significance (SKAT-O *P*≤4.37×10^-5^) versus all non-significant genes (*n*=60,605). Error bars indicate 95% CIs as defined by +/− 1.96 standard errors of the mean normalised mRNA count. **b,** Selection strategy for genes to include in conditional knockdown experiment. Numbers in parentheses indicate the number of genes in a given group. **c,** Experimental design for CRISPR knockdown, adipocyte cell culture and imaging. **d,** Confirmation of knockdown efficacy using relative mRNA expression in differentiated adipocytes. Error bars indicate 95% CIs defined by +/− 1.96 standard error of the mean. One-sided Wald test comparing mRNA count in knockdown cell line relative to count in Cas9 empty, with the alternative hypothesis that the count is lower in the knockdown: \*\*\**P*<0.001, \*\**P*<0.01, \**P*<0.05. **e,** Confirmation of knockdown efficacy using relative RNA expression quantified with qPCR in undifferentiated adipocytes. Error bars indicate 95% CIs defined by +/− 1.96 standard error of the mean. One-sided t-test comparing mRNA count in knockdown cell line relative to count in Cas9 empty, with the alternative hypothesis that the count is lower in the knockdown: \*\*\**P*<0.001, \*\**P*<0.01, \**P*<0.05. Knockdown confirmation for *PPARG* was performed by checking two isoforms, *PPARG1* and *PPARG2*. ^†^*PPARG* knockdown confirmation was performed in differentiated adipocytes because expression of *PPARG* isoforms was too low in undifferentiated adipocytes.

We selected four guide RNAs per gene target to generate 14 knockdown hWAT cell lines (Methods, Figure 3c, and Supp. Table 9). We additionally generated a negative control cell line with Cas9 empty vector. We confirmed gene knockdown through a two-step process: (1) RNA-seq expression of the target gene in knockdown cell lines relative to expression in Cas9-empty cell line and observed significantly decreased (*P*<0.05, one-sided Wald test) expression for *COL5A3, DENND5B, EXOC7, HERC1, INSR, MFAP5, MLXIPL, SLTM, TRIP10*, and *UBR2* (Figure 3d, Supp. Table 10) and (2) qPCR measurements of RNA for genes whose knockdown could not be confirmed via RNA-seq and observed significantly decreased (*P*<0.05, one-sided *t*-test) for *ABCA1* and *PPARG* (Figure 3e, Supp. Table 12). Expression of *PCSK1* quantified by qPCR was not significantly decreased compared to the control (qPCR fold change [95% CI]=0.624 [0.373, 1.045], one-sided *t*-test *P*=0.0665,), but was suggestive of a knockdown. We retained the *PCSK1* possible knockdown in downstream analysis, with a note of caution in the interpretation of results. The only gene knockdown which could not be confirmed and was not close to having significantly decreased expression was *PLIN1* (qPCR fold change [95% CI]=0.933 [0.390, 2.234], one-sided *t*-test *P*=0.450).

Abnormal adipogenesis and lipid accumulation are hallmarks of adipose tissue dysfunction^45^. As the process of adipocyte differentiation is considered a necessary precursor to lipid accumulation^46^, we use BODIPY staining^47^ (Methods) to measure both lipid accumulation and adipogenesis more generally. The BODIPY assay was conducted for six replicates of each knockdown cell line.

### Knockdown of obesity and fat distribution associated genes alters *in vitro* adipogenesis and lipid accumulation

We observed significantly reduced (*P*<0.05 in two-sided *t*-test of mean fold change [FC] in fluorescence between knockdown and control Cas9-empty cell line) lipid accumulation in the *PPARG* knockdown cell line (Figure 4, Supp. Table 13). Inactivation of *PPARG* through knockout has previously been shown to lower lipid accumulation in human and mouse pre-adipocytes^29,48,49^, providing validation of the assay used here.

**Figure 4.**
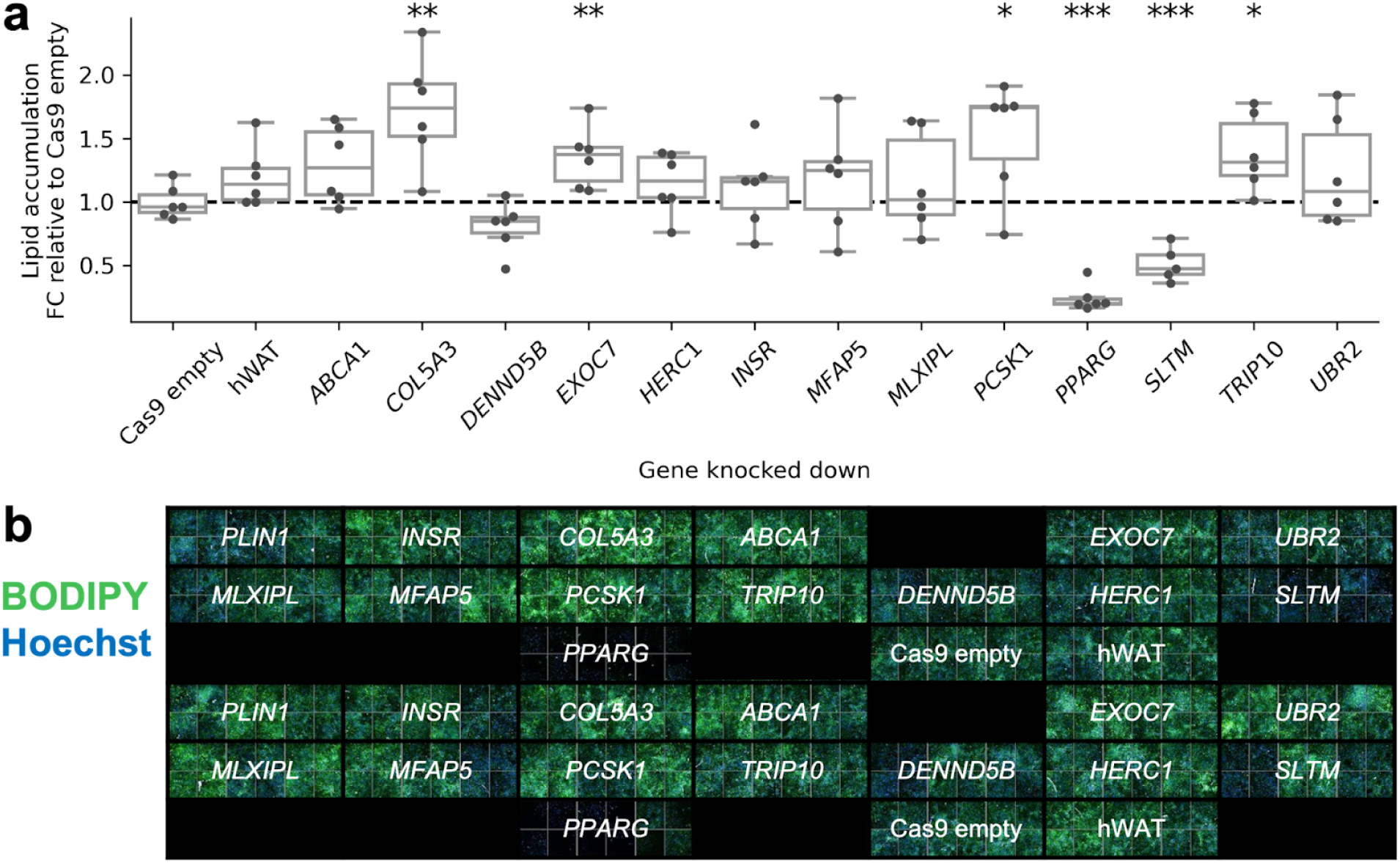
Effect of CRISPR single-gene knockdown on lipid accumulation in human adipocytes. **a**, Effect of single-gene knockdown on lipid accumulation measured by whole-well mean cytoplasm intensity of BODIPY dye fluorescence. Points represent measurements from separately-cultured cells (biological replicates) from the same knockdown cell line. Each knockdown cell line has six replicates, except *SLTM*, which had one outlying replicate removed (Methods). **b,** Differentiated human adipocyte knockdown cell lines stained with BODIPY (green) and Hoechst (blue), imaged at 10x magnification. The elements of the box plots are median (horizontal line in box), upper (Q3) and lower (Q1) quartiles marking the ends of the box, with whiskers extending to the most extreme points within the range [Q1-IQR, Q3+IQR], where IQR=Q3-Q1. All points are shown. Two-sided *t*-test of mean fold change relative to Cas9-empty cells: \*\*\**P*<0.001, \*\**P*<0.01, \**P*<0.05. Summary statistics are available in Supp. Table 13. FC, fold change. hWAT, human white adipose tissue.

We also noted reduced lipid accumulation in knockdown of the novel target *SLTM* (FC=0.514, *P*=1.91×10^-4^) (Figure 4a, Supp. Table 13). *SLTM* encodes the SAFB-like transcription modulator, which regulates the GLI family of transcription factors in mice^50^. These GLI transcription factors in turn control expression of lipid metabolic genes, including critical adipogenesis transcription factors *PPARG* and *C/EBP* (α, β, γ, and δ)^51^. Notably, the *SLTM* pLoF burden association was replicated in a multiancestry meta-analysis conducted in All of Us^52^ as the top association for BMI (All of Us meta-analysed *P*=2.75×10^-7^). While this suggests a novel therapeutic target for altering fat deposition, more work is needed to understand why individuals carrying rare pLoF variants in *SLTM* have higher BMI (burden beta (SE) = 0.0191 (0.00455), SKAT-O *P*=2.65×10^-6^), and the target tractability of SLTM, which may be amenable to small molecule binding (Supp. Table 14).

We observed significantly increased lipid accumulation relative to Cas9-empty cells in knockdowns of *EXOC7* (FC=1.35, *P*=0.0096), *TRIP10* (FC=1.39, *P*=0.0157), and *COL5A3* (FC=1.72, *P*=0.0028) (Figure 4a, Supp. Table 13). The products of these genes are all variously involved in lipid uptake, synthesis, and adipogenesis. EXOC7 is a component of the exocyst complex, which regulates the uptake of free fatty acids by adipocytes^53^. TRIP10 is an interactor of the thyroid hormone receptor (TR-β1)^54^, which regulates *de novo* fatty acid synthesis^55^. TR-β1 has been proposed as a target to treat dyslipidaemia^56^, and the TR-β1 agonist resmetirom is currently in Phase III clinical trials for treating non-alcoholic fatty liver disease^57^. However, there are no known drugs directly targeting TRIP10 (Supp. Table 14). We also observed increased lipid accumulation in knockdowns of *COL5A3* (FC=1.72, *P*=2.78×10^-3^) (Figure 4a, Supp. Table 13) – as a ubiquitous component of the extracellular matrix, type V collagen informs the proper differentiation and development of adipocytes^58^. A Phase II clinical trial of collagenase histolyticum, a hydrolytic enzyme that degrades COL5A3, has proven successful in dissolving lipomas, i.e. benign lumps of fatty tissue underneath the skin^59^ (Supp. Table 14). The *PCSK1* knockdown cell line also demonstrated significantly increased lipid accumulation, however we could not draw any conclusions because the knockdown was not confirmed.

### Transcriptome-wide effects of gene knockdowns

Obesity-gene knockdowns had wide-ranging effects on the hWAT transcriptome, with between eight and 1,904 genes differentially expressed (|log_2_(fold change)|>1, FDR-adjusted *P*<0.05) in knockdown cell lines as compared to Cas9-empty negative controls (Figure 5 and Supp. Table 15). We examined the molecular effects of these knockdowns through pathway enrichment analyses for pathways in the KEGG^60^, REACTOME^61^, and GO:Biological Process and GO:Molecular Function^62^ databases.

**Figure 5.**
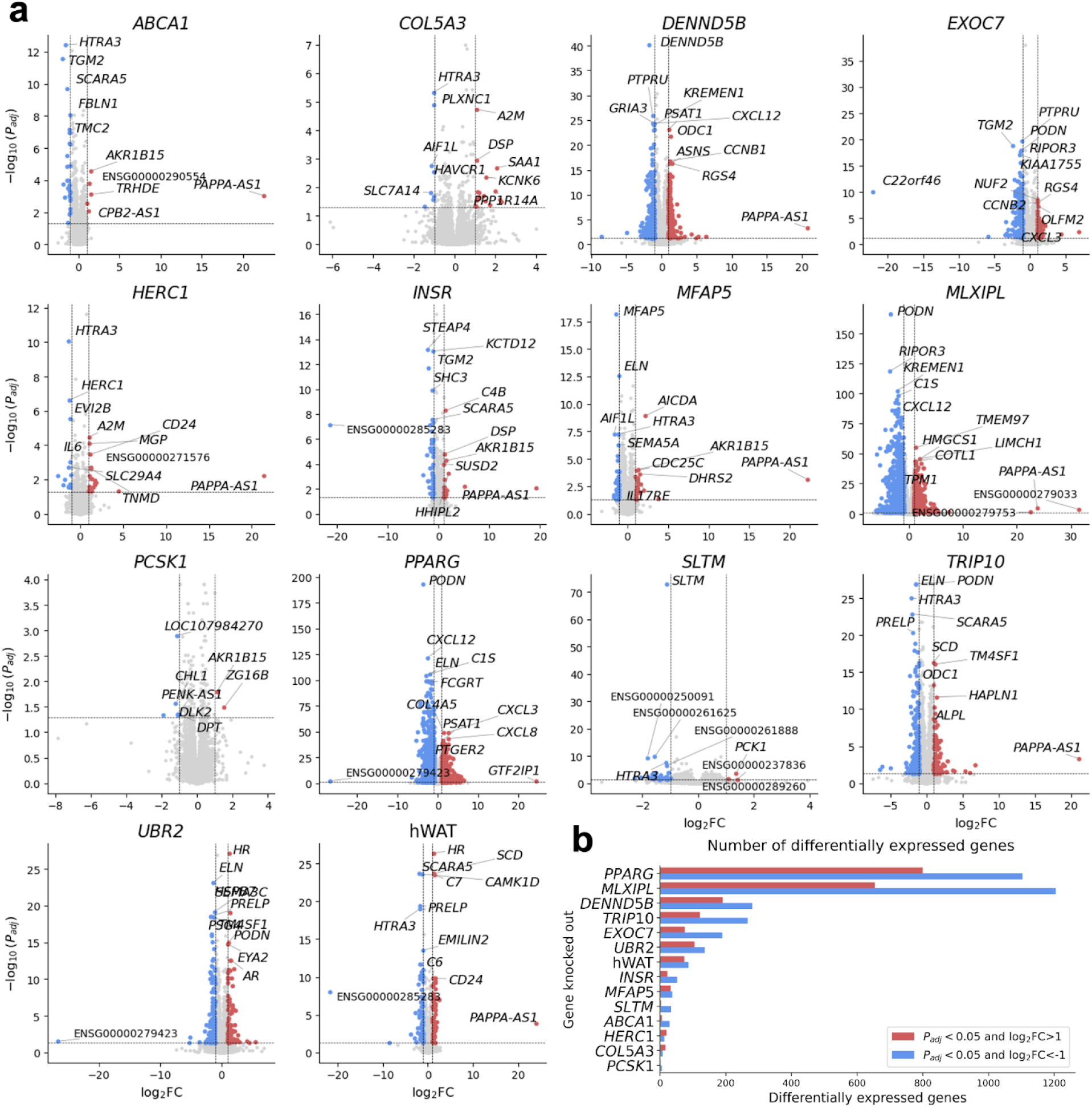
Effect of gene knockdown on differential gene expression in human adipocytes. **a**, Gene differential expression results from RNA sequencing data performed on hWAT wild-type and single-gene knockdowns. Gene differential expression analysis was performed with DESeq2^70^, comparing each cell line (wild-type hWAT or knockdown) to the Cas9-empty cell line. All significance thresholds use *P*_adj_, a *P*-value corrected for multiple testing calculated in DESeq2. Blue points indicate genes which are significantly downregulated (*P*_adj_<0.05, log_2_FC<1). Red points indicate genes which are significantly upregulated (*P*_adj_<0.05, log_2_FC>1). Grey points are all other genes. Genes are labelled if they are in the top five most significant (lowest *P*_adj_) red or top five most significant blue points, in the top five most significant regardless of point colour, or have abs(log_2_FC)>10. The horizontal and vertical dashed lines indicate the significance cutoffs of *P*_adj_<0.05 and abs(log_2_FC)>1, respectively. **b,** Counts of significantly downregulated (blue, *P*_adj_<0.05 and log_2_FC<-1) and upregulated (red, *P*_adj_<0.05 and log_2_FC>1) genes for each single-gene knockdown and hWAT. The y-axis is sorted by the total number of differentially expressed genes for each knockdown condition. FC, fold change.

Knockdown of *PPARG*, a master regulator of adipogenesis, significantly alters 95 pathways (Family-wise error rate [FWER] *P*<0.05), including: (1) transcriptional regulation through PRC2-driven DNA methylation (number of genes (*n*)=59, *P*<0.0001), histone acetylation (*n*=138, *P*<0.0001), and small RNAs (*n*=102, *P*=0.0001), (2) protein synthesis by modulating rRNA expression through SIRT1 (*n*=63, *P*<0.0001), ERCC6 (CSB) and EHMT2 (G9a) (*n*=71, *P*<0.0001) and the B-WICH chromatin remodelling complex (*n*=86, *P*=0.0001), and (3) functions critical to cell division such as assembly of the ORC complex at the origin of replication (*n*=64, *P*<0.0001), condensation of prophase chromosomes (*n*=69, *P*<0.0001), and cell cycle checkpoints (*n*=286, *P*<0.0001) (Supp. Table 16). As expected, *PPARG* knockdown also alters pathways important for adipogenesis, such as the transcription of androgen-receptor regulated genes (*n*=62, *P*<0.0001), RUNX1 regulation^63^ (*n*=89, *P*=0.0001), formation of the beta-catenin TCF trans-activating complex^64^ (*n*=87, *P*=0.0001), and pre-NOTCH expression and processing^65^ (*n*=104, *P*=0.0003) (Supp. Table 16).

We observed enrichment of several mitotic, transcription, and translation pathways in knockdowns of lipid accumulation-altering genes *EXOC7* and *TRIP10* (Supp. Table 16), but did not find mechanisms directly implicated in lipid accumulation. On the other hand, while *MLXIPL* knockdown did not affect lipid accumulation in our assays, we nevertheless observed enrichment (FWER *P*<0.05) of pathways involved in adipogenesis, such as interferon α/β signalling^66^ (*n*=66, *P*=0.0016) and lipopolysaccharide binding^67^ (*n*=32, *P*=0.0284). *MLXIPL* encodes the carbohydrate-responsive element-binding protein (ChREBP), which is thought to regulate gene expression in response to glucose and reduces adipose tissue mass in mouse knockout models^68^. While this may be a potential therapeutic target for fat distribution, mice deficient in MLXIPL/ChREBP do not tolerate fructose in their diet and develop severe diarrhoea and irritable bowel syndrome^69^.

## Discussion

Through a series of population-level genetic and *in vitro* functional genomics investigations, we demonstrate a model to map the biological mechanisms of obesity and body fat distribution, from variant to molecular function to systemic phenotype. We utilised complementary rare variant gene-based analyses and common variant prioritisation to nominate 69 genes with robust associations to BMI, WHRadjBMI, body fat percentage, and six tissue-derived fat components in up to 402,375 participants in UKB. By evaluating the sex-specific and phenome-wide associations of these genes, we built a comprehensive understanding of their systemic effects. Combining multiple lines of genomic, transcriptomic, and prior functional evidence, we selected 14 genes for functional characterisation by CRISPR knockdown in human white adipose tissue (hWAT) cell lines. Given the difficulty of manipulating adipocytes in high-throughput experiments, we believe this is one of the first studies to comprehensively identify and target multiple genes for functional characterisation in adipogenesis. We observed reduced lipid accumulation in *PPARG* and *SLTM* knockdown, and increased lipid accumulation in knockdowns of *COL5A3, EXOC7* and *TRIP10*. While some of these genes have previously been identified through large-scale genetic analyses, we show their functional effects on adipogenesis for the first time, and highlight potential therapeutic avenues to regulate body fat mass and/or distribution. Taken together, our population-based and *in vitro* genetics investigations highlight molecular mechanisms of, and therapeutic avenues to alter, fat distribution.

The recent growth in sample sizes of whole-exome sequenced participants in biobanks has accelerated the development of gene-level analyses^71–73^. By annotating rare variants with their putative functional consequences and then collapsing these across each gene, gene-level testing has enabled the discovery of novel genes associated with complex human traits^42,74^. While gene-level testing is a powerful strategy for gene discovery, it remains limited by the requirement of high-quality functional annotations of rare variants. Here, we combined evidence from CADD, REVEL, and SpliceAI annotations^35–37^ to generate high-confidence sets of variants, and integrated evidence from common variant studies to prioritise putative risk genes for functional follow up. Using these annotation categories, we built allelic series that showed the effect of each category of variants within a given gene. In assessing the monotonicity of these allelic series, we made the simplifying assumption that missense variants not predicted to be LoF or highly damaging would act in the same direction as pLoFs or damaging variants. However, missense variants may also result in gain of function^75^. Categorising missense variants into gain– and loss-of-function before assessing monotonicity could result in identifying more genes with monotonic allelic series. Furthermore, our analysis was also only focused on coding variation. In the future, improved rare variant masks, such as those that better stratify missense variants or incorporate non-coding rare variation outside exonic regions, may help identify additional therapeutic targets to alter fat distribution.

Genome-wide common variant and exome-wide rare variant analyses can provide complementary lines of evidence to nominate genes associated with human traits^74,76^. We found that genes implicated for obesity and body fat distribution by a burden of rare damaging variation were: (1) associated with rare Mendelian forms of obesity and lipodystrophies, and (2) also prioritised by common variant prioritisation scores^38^ for BMI, WRHadjBMI and body fat percentage. Converging evidence from across the allele frequency and phenotypic severity spectra can therefore identify high-confidence genes for a range of human traits. Assessing the sex-specific and phenome-wide consequences of these genes can provide evidence for therapeutic potential, for example by revealing possible side-effects on platelet distribution or neutrophil percentage.

We measured the effects of CRISPR gene knockdowns on lipid accumulation in hWAT cell lines using the BODIPY assay, which we performed in six technical replicates per knockdown cell line to ensure replicability. As the process of adipocyte differentiation is considered a necessary precursor to lipid accumulation^46^, we interpreted the BODIPY readout as a measure of both lipid accumulation and adipogenesis more generally. However, we acknowledge that lipid accumulation is only one of many pathways involved in adipocyte behaviour, alongside glucose uptake, stem cell dynamics, adipocyte lifespan, depot-specific effects, etc. Thus, a failure to find alterations in lipid accumulation and adipogenesis due to gene knockdown *in vitro* does not imply that the gene is not relevant for obesity nor that it is an unattractive drug target. Genetic variation associated with obesity will likely affect other components of the adipose tissue niche, including innervation, immune cells, or stromal components in a depot-specific manner^77–79^. Indeed, BMI-associated genetic variation is known to alter the expression of genes in the central nervous system^12^, and blockbuster GLP1-receptor agonists that are prescribed for weight loss^13,14^ act primarily through the dorsomedial hypothalamus to control food intake^15^. Therapeutic targets against obesity will thus need to span the range of mechanisms through which excess body fat may accumulate.

Our confirmed knockdown of *INSR* did not significantly affect lipid accumulation as measured by the BODIPY assay, despite this gene having a generally well-characterised role in body fat accumulation^80,81^. However, *INSR* was still partly expressed (RNA-seq expression fold change (FC) [95% CI]=0.521 [0.452, 0.601]), which may explain why the knockdown did not result in significant changes to lipid accumulation.

Of the two genes carrying rare missense variation associated with increased BMI and with confirmed knockdowns, *EXOC7* knockdown increased lipid accumulation in hWAT cell lines while *SLTM* knockdown decreased lipid accumulation. Rare missense variation in *PPARG* is associated with decreased body fat percentage and the disruption of *PPARG* in hWAT cell lines resulted in decreased lipid accumulation. Similarly, of the two genes affecting central obesity (WHRadjBMI or ratio of android to gynoid tissue fat percentage) with confirmed knockdowns significantly increasing lipid accumulation, *COL5A3* was correspondingly associated with increased central obesity, whereas *TRIP10* was associated with decreased central obesity. Our results demonstrate the importance of systematically characterising the functional effects of gene perturbation at the level of cell lines and model organisms, which may not follow the expected direction of effect based on missense variation in humans.

In summary, we demonstrate that the convergence of evidence across rare and common genetic variation can help identify high-confidence target genes for overall adiposity and body fat distribution. We provided in-depth functional characterisation through CRISPR knockdowns in human adipocytes, allowing us to nominate candidate genes for therapeutic targets. Through functional readouts and transcriptomic analyses, we also highlighted several novel molecular mechanisms by which genetic variation may impact adipogenesis. Our results provide a model by which future work can integrate genetic and functional evidence to identify, design, and validate potential drug targets to alter overall and tissue-specific fat distribution.

## Methods

### Imputed data quality control

#### Sample quality control and assigning population labels

Beginning with the 487,409 individuals with phased and imputed genotype data, we restricted to unrelated individuals with low autosomal missingness rates used for PCs by Bycroft *et al*.^82^. We then used genotyping array data, subset to LD-pruned autosomal variants, from these samples to project into the PC space defined by the 1000 Genomes dataset^83^, ensuring that we correctly account for shrinkage bias in the projection^84^. Next, we used the ‘super-population’ labels (AFR=Africans, AMR=Admixed Americans, EAS=East Asians, Europeans=EUR, South Asians=SAS) of the 1000 Genomes dataset to train a RF classifier, using the randomForest (4.6) library in R^85^, and predicted the super-population for each of the UKB samples. Samples with classification probability >0.99 for the European super-population, were retained for downstream analysis.

Following our ancestry based filtering regime, we remove samples who withdrew from UKB participation as well as those individuals who were omitted from phasing and imputation. For GWAS, we further subset individuals passing imputed data quality control to those who also passed exome sequencing quality control (described below), resulting in a set of 397,315 individuals.

#### Variant quality control

Over 92 million imputed variants across the autosomes and chromosome X are available for analysis. As a starting point for our initial collection of GWAS, we subset to variants with MAF >0.1% in the subset of individuals defined in the sample QC procedure and an INFO score >0.8 from the UKB SNP manifest file. Following this collection of filtering steps, 16.7 million variants were retained for common variant GWAS. After performing GWAS, two additional filters were performed, retaining variants with Hardy-Weinberg *P*>1×10^-10^, calculated on the whole QCed subset of individuals, and retaining variant-level summary statistics if MAF >0.1%, calculated for each GWAS. This results in 13,117,850 variants for downstream common variant analysis.

### Exome sequencing data quality control

#### Exome sequencing data quality control summary

Using the UKB Research Access Platform we accessed the gnomAD VCFs containing GATK-called genotypes for 454,671 individuals. We followed an identical sample QC protocol to Karczewski *et al.*^42^, filtering individuals on covariate-regressed individual-level metrics using median absolute deviations. We removed variants flagged by the gnomAD QC team for failing one or more of their filters (*AC0*, *RF*, *MonoAllelic*, *InbreedingCoeff*). Genotype calls were set to missing if they failed filters for genotype quality, depth and alelle balance (described in Karczewski *et al.*^42^). We filter to European ancestry using the super-population ancestry labels assigned with an RF classifier, described above for imputed data quality control.

#### Variant and sample-level QC

We defined ‘high quality’ variants as those MAF>0.1% and call rate ≥0.99 falling within the UKB capture intervals plus 50 bp padding. These variants were used to evaluate sample-level metrics of mean call rate and depth and retained samples satisfying all of the following:

● Genetic sex inferred as XX or XY (specifically, genetic sex is defined).
● Mean call rate ≥0.99 among high quality variants.
● Mean coverage ≥20x among high quality variants.
● Not withdrawn.

Next, we removed variants satisfying at least one of the following criteria:

● The variant lies outside the UKB capture plus 50 bp padding.
● The variant lies within a low complexity region.
● The variant lies within a segmental duplication.

Among this (sample, variant) set, we ran Hail’s `sample_qc()`^86^ to remove samples lying outside the median±4 median absolute deviations (MADs) within each super-population (see above section on imputed data quality control). The QC protocol was split by UKB ES tranche (50k, 200k, 450k) to guard against batch effects, as tranches were sequenced in separate runs. The following metrics were used for QC:

● Number of deletions (`n_deletion`).
● Number of insertions (`n_insertion`).
● Number of SNPs (`n_snp`).
● Ratio of insertions to deletions (`r_insertion_deletion`).
● Ratio of transitions to transversions (`r_ti_tv`).
● Ratio of heterozygous variants to homozygous alternate variants (`r_het_hom_var`).

Following MAD filtering (Supp. Figure 1, Supp. Table 1), 402,375 European samples were retained for analysis. For each sample, we excluded non-passing sites as described in Karzcewski *et al.*^42^. Briefly, an RF classifier was trained to distinguish true positives from false positive variants using a collection of allele and site annotations. Variants were assigned ‘PASS’ to maximise sensitivity and specificity across a series of readouts in trio data and precision-recall in two truth samples, after which samples with excess heterozygosity (defined as inbreeding coefficient <-0.3) were removed. Next, we removed low quality genotypes by filtering to the subset of genotypes with depth ≥10 (5 among haploid calls), genotype quality ≥20, and minor allele balance >0.2 for all alternate alleles for heterozygous genotypes. Following this filter, we remove variants that were not called as ‘high quality’ among any sample. The resulting high quality European call set consisted of 402,375 samples and 25,229,669 variants.

### Variant consequence annotation

We annotated exome-sequencing variants using Variants Effect Predictor (VEP) v105 (corresponding to GENCODE v39)^87^ with the LOFTEE v1.04_GRCh38^34^ and dbNSFP^88^ plugins, annotating variants with CADD v1.6^35^, and REVEL using dbNSFP4.3^36^ and loss-of-function confidence using LOFTEE. We provide code and instructions for this step in our VEP_105_LOFTEE repository^89^, which contains a Docker/Singularity container for reproducibility of annotations. Next, we ran SpliceAI v1.3^37^ using the GENCODE v39 gene annotation file to ensure alignment between VEP and SpliceAI transcript annotations. For variant-specific annotations we use ‘canonical’ transcripts. We separated variants by transcript using `bcftools +split-vep` and filtered to MANE Select^90^ protein-coding transcripts. If the gene lacked a MANE Select transcript we selected the canonical transcript defined by GENCODE v39. Using this collection of missense, pLoF, and splice metrics, and annotations of variant consequence on the canonical transcript, we then determine a set of variant categories for gene-based testing.

#### Variant consequence categories

1. **High confidence pLoF**: High-confidence LoF variants, as defined by LOFTEE^34^ (LOFTEE HC).
2. **Damaging missense/protein-altering**: At least one of:
  a. Variant annotated as missense/start-loss/stop-loss/in-frame indel and (REVEL≥0.773 or CADD≥28.1 or both).
  b. Any variant with SpliceAI delta score (DS)≥0.2 where SpliceAI DS the maximum of the set {DS_AG, DS_AL, DS_DG, DS_DL} for each annotated variant (where DS_AG, DS_AL, DS_DG and DS_DL are delta score (acceptor gain), delta score (acceptor loss), delta score (donor gain), and delta score (donor loss), respectively).
  c. Low-confidence LoF variants, as defined by LOFTEE (LOFTEE LC)
3. **Other missense/protein-altering**: Missense/start-loss/stop-loss/in-frame indel not categorised in (2).
4. **Synonymous**: Synonymous variants with SpliceAI DS<0.2 in the gene.

REVEL and CADD score cut-offs are chosen to reflect the supporting level for pathogenicity (PP3) from the American College of Medical Genetics and Genomics and the Association for Molecular Pathology (ACMG/AMP) criteria^91^.

### Phenotype curation

We used the following nine phenotypes, either directly measured by UKB or derived from UKB phenotypes: body mass index (UKB code: 21001), waist-to-hip ratio (derived from waist circumference [UKB code: 48] and hip circumference [UKB code: 49]) with BMI regressed out, body fat percentage (UKB code: 23099), android tissue fat percentage (UKB code: 23247), gynoid tissue fat percentage (UKB code: 23264), ratio of android to gynoid tissue fat percentage (derived from android and gynoid tissue fat percentage phenotypes), total tissue fat percentage (UKB code: 23281), visceral adipose tissue volume (UKB code: 22407), abdominal fat ratio (UKB code: 22434).

### Genetic association testing

#### Genetic association testing summary

All variant– and gene-level associations were performed in the European-ancestry subset of the UKB using the Scalable and Accurate Implementation of GEneralised mixed model (SAIGE)^71^, a mixed model framework that accounts for sample relatedness.

In SAIGE step 0, we constructed a genetic relatedness matrix (GRM) using the UKB genotyping array data. The genotyped data is LD pruned using PLINK (`--indep-pairwise 50 5 0.05`) ^92^, and the sparse GRM is calculated using the `createSparseGRM.R` function within SAIGE, using 5,000 randomly selected markers, with relatedness cutoff of 0.05,

To generate a variance ratio file for subsequent steps in SAIGE, we selected 2000 variants from the genotyping array data to define a PLINK dataset. For testing common variants in imputed data, we extracted 2000 variants with MAC≥20. For testing the exome-sequencing data, we extracted two sets of 1000 variants with 10≤MAC<20 and MAC≥20, and combined these sets of markers.

In SAIGE step 1 for each trait, the null model is fit using the curated phenotype data and sparse GRM, with no genetic contribution. Default parameters were used in SAIGE, except `--relatednessCutoff 0.05`, `--useSparseGRMtoFitNULL TRUE` and `--isCateVarianceRatio TRUE.` In the sex-combined analyses, we account for *age, sex, age*^2^*, age × sex, age*^2^ *× sex*, the first 21 PCs, UKB assessment centre, and exome-sequencing tranche (50k^93^, 150k^94^ or 250k^95^ tranches) as fixed effects. For sex-specific analyses, we exclude sex-dependent terms (*sex, age × sex, age*^2^ *× sex*) when accounting for fixed effects. All continuous traits were inverse rank normalised using the `--invNormalize` flag in SAIGE. For SAIGE step 2, we always use the flag `--LOCO FALSE` and –`-is_fastTest TRUE`.

#### Common variant association testing in imputed data

We performed single-variant genetic association testing of the UKB imputed data using SAIGE version 1.1.6.3 on the Oxford Biomedical Research Computing cluster. For consistency across analyses, only individuals who passed sample-level quality control in both the imputed and exome-sequencing data were included.

Following null model fitting, we carried out variant testing in SAIGE step 2 with default parameters, except for –-is_Firth_beta TRUE and –-pCutoffforFirth=0.1.

##### Finemapping

Using summary statistics from the common variant association tests, finemapping loci are identified for each trait. Starting with the most significant variant in the pool of genome-wide significant variants, a 1 Mb window centred around the variant is created. All genome-wide variants falling in this window are considered part of the locus defined by the most significant variant and removed from the pool of genome-wide significant variants. Then we proceed to the next most significant variant in the pool of genome-wide significant variants and repeat until the pool of genome-wide significant variants is empty. Loci with overlapping windows are merged.

LD matrices were calculated with LDstore v2.0^96^, using the same set of individuals used in the GWAS for each trait. Finemapping was performed with FINEMAP v1.4^96^, using shotgun stochastic search and allowing a maximum of 10 causal SNPs per locus.

#### Rare variant and gene-level testing in exome sequencing data

We carried out rare variant and gene-level genetic association testing in the European-ancestry subset of the UKB exome sequencing data using SAIGE. All analyses involving the exome sequencing data was carried out on the UKB Research Analysis Platform (RAP) using SAIGE version wzhou88/saige:1.1.9^71^.

Following null model fitting, we carried out variant and gene-level testing in SAIGE step 2 using the variant categories described above, with the `--is_single_in_groupTest TRUE` flag. All other parameters were set to default, except `--maxMAF_in_groupTest=0.0001,0.001,0.01`, although we only report results for maximum MAF of 0.01. We included the following collection of group tests, using the annotations defined above (see ‘Variant consequence annotation’ methods section):

● High confidence pLoF
● High confidence pLoF or damaging missense/protein-altering

For genes with no damaging missense/protein altering variants, only the pLoF variant consequence mask was considered when performing Bonferroni multiple-testing correction. Variant-level and gene burden tests are always two-sided.

##### Obesity and fat distribution trait gene-level association tests

For each gene and for each obesity and fat distribution trait we tested the SKAT-O association of rare variant (MAF<1%) variation. Exome-wide statistical significance (*P* < 1.58×10^-7^) for gene-level tests was defined using Bonferroni correction for 315,996 unique phenotype-gene-consequence mask combinations. To estimate the strength and direction of effect of damaging variation in genes on phenotypes, we also performed gene burden tests, however, the *P*-values from the burden analysis were not used to determine significance.

To expand the set of obesity and fat distribution associated genes to consider for functional screening, we followed an FDR-control process similar to Zhou *et al.*^97^. We selected the gene-level association result which had the lowest SKAT-O *P*-value (across all phenotypes and both variant masks) and applied the Benjamini-Hochberg procedure to identify 83 significant genes when controlling FDR to 1% (corresponding to a *P*-value of 4.37×10^-5^). If we had performed the FDR-controlled selection using the full set of results instead of taking the result with the lowest *P*-value this would be equivalent to an FDR of 12.0%.

We re-ran the gene-level association tests for the 83 FDR-significant genes, conditional on the top finemapped common (MAF>0.1%) variants for finemapped loci (described above). For each trait, we identified finemapped common variants for each trait which have the highest Bayes factor for being a causal variant (i.e. strongest evidence for causality) in their respective loci. Variants tied for the highest Bayes factor are all selected. There are a total of 915 finemapped trait-variant pairs (846 unique variants) on which to condition. We used the `--condition` flag in SAIGE to condition on the selected common variants when performing gene-level tests for the corresponding phenotype. We perform the gene-level association tests for each chromosome independently and only conditioned on common variants from the same chromosome as the genes being tested.

After conditioning, seven genes (*ACVR1C, PRPH, PYGM, SEC16B, TNFRSF6B, TRIM40, TUBE1*) were no longer significant at the predefined SKAT-O *P*≤4.37×10^-5^ threshold, leaving 76 genes. Of these 76, seven genes (*DEFB112*, *CHMP4B*, *FEZF2*, *GLP1R*, *PCBD2, TM4SF20, VGF*) were flagged as having very low minor allele count (MAC<10), leaving 69 genes to be considered for functional screening.

#### Common variant gene prioritisation scores

We used Polygenic Priority Scores (PoPS) calculated by Weeks *et al.*^38^ for UKB BMI, WHRadjBMI and body fat percentage. Scores were downloaded from https://www.finucanelab.org/data. There were 34,635 gene-level PoPS available each for BMI, WHRadjBMI and body fat percentage, totalling 103,905 gene-trait PoPS.

#### Sex difference analyses

We use sex-specific estimated burden effect sizes (β) and standard errors (SE) to calculate *z*-scores corresponding to sex-differential effects using the heterogeneity test described by Martin *et al.*^98^:

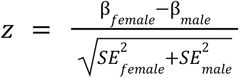

The sex-differential *P* is estimated under a two-sided hypothesis:

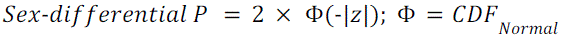

Genes were considered to exhibit significant sex-differential effects if *P*<2.67×10^-6^, Bonferroni adjusted for 18,737 genes tested for sex-differential effects.

#### Age at diagnosis analyses

We curated age at diagnosis of obesity from the UKB linked primary care and hospital record data using mapping tables generated by Kuan *et al.*^99^. Any codes related to “history of obesity”, for which accurate age at diagnosis could not be extracted, were excluded. We left-truncated observations at the age of the first record (of any code) in either primary care or hospital data, and right-censored at the age of the last record. For each of the 69 FDR-significant genes identified from ES analyses, we performed Cox-proportional hazards modelling^100^ to estimate differences in lifetime risk of developing obesity between carriers of pLoF, damaging missense, other missense, synonymous, or non-coding variants (at MAF<1%) and wild-type individuals (reference group). All effects were adjusted for sex (in sex-combined analyses), the first 10 genetic PCs, birth cohort (in ten-year intervals from 1930 to 1970), and UKB assessment centre. Cox modelling was performed using the R package survival v3.3.1^101^. We visualised age at onset probabilities using Kaplan-Meier survival curves with the R package survminer v0.4.9^102^.

#### Genebass gene-level summary statistics

Gene-level association test summary statistics were calculated by Karczewski *et al.*^42^ and downloaded as a Hail Matrix Table from https://app.genebass.org/downloads. Only pLoF-associated results corresponding to our 69 FDR≤1% genes were used. Associations were significant if Genebass SKAT-O *P*≤9.98×10^-6^, controlled for FDR≤1% using the Benjamini-Hochberg procedure^39^.

#### Genetic correlation estimates

Genetic correlation estimates were accessed from the Neale lab’s UKB genetic correlation browser: https://ukbb-rg.hail.is/rg_browser/^44^. These genetic correlations were estimated using LD Score regression^43^ v1.0.1, using summary statistics from common (MAF>0.1%) imputed variant GWAS. Genetic correlation *P*-values less than or equal to 1×10^-308^ are reported in Supp. Table 5 as zero due to numerical precision limits in Microsoft Excel.

#### Pathway enrichment

##### CRISPR RNA sequencing

We used GSEA^103^ v4.3.3 to test for pathway enrichment in CRISPR adipocytes RNA sequencing data. Regularised log transformed mRNA counts calculated by DESeq2 are provided to GSEA. The pathway enrichment for each knockdown or wild-type hWAT is compared against the baseline control, the empty Cas9 vector, such that pathways with more positive normalised enrichment scores (NES) are those enriched in Cas9 empty and those with more negative NES are those enriched in the knockdown or hWAT.

We used pathway sets from KEGG Legacy, KEGG Medicus, REACTOME, GO:Biological Process, and GO:Molecular Function, downloaded from GSEA’s Molecular Signatures Database v2023.2.Hs^103^. We always used 10,000 permutations when running GSEA.

#### Selecting target genes

To choose target genes for CRISPR-Cas9 knockdown we consider all MAC≥10 gene association results with FDR<0.01 which remain significant (*P*≤4.37×10^-5^) after conditioning on common variation significantly associated with the trait (Methods). We then compiled multiple lines of evidence for each gene:

● Sufficiently expressed in wild-type hWAT cells (RNA-seq transcript counts >10 at eight and 24 days after differentiation).
● Monotonic allelic series indicating a dose-response relationship with gene dosage and a obesity or fat distribution trait.
● Significant obesity age-of-onset association.
● Published functional work involving gene knockout or knockdown implicates the role of the gene in adiposity.

Using these lines of evidence we separated the genes into categories: positive controls (genes previously implicated in obesity pathways by functional work in human adipocytes), plausible candidates (genes which have not been implicated in adiposity pathways by functional work in human adipocytes, but have suggestive evidence from other functional work or from GWAS/burden association results), and impossible candidates (RNA-seq counts are too low or gene seems to be associated with a pathway that we are not testing). From these categories we chose 14 target genes for knockdown (*ABCA1*, *COL5A3*, *DENND5B, EXOC7*, *HERC1, INSR*, *MFAP5*, *MLXIPL*, *PCSK1*, *PLIN1, PPARG, SLTM, TRIP10, UBR2*).

#### Cell culture and reagents

Human white adipose tissue cells were cultured in Dulbecco′s Modified Eagle′s Medium (DMEM, Sigma Aldrich, Cat #D6546) with 10% foetal bovine serum (FBS, ThermoFisher Cat # A5256801) in a 37°C humidified incubator with 5% CO_2_^104^. Passage 40 was used for the experiments.

#### Cell line generation

For each gene target, four guide RNAs (gRNAs) were commercially synthesised and cloned into the lentiviral vector pLentiCRISPRv2 (GenScript, UK). gRNA sequences are listed in Supp. Table 9. Lentivirus for individual gRNAs was produced. Briefly, CRISPR plasmids were cotransfected in HEK293T with packaging vectors pMD2.G (Addgene, 12259) and psPAX2 (Addgene, 12260) using FuGENE® HD Transfection Reagent (Promega Cat #E2311). Viral supernatant was collected at 48 hours and 72 hours post-transfection for each gRNA. The lentivirus from the four gRNAs were pooled. hWAT cells were transduced with the pooled virus and with 2 µg/ml polybrene. The selection was performed for 3 days in 1 μg/ml puromycin with media changes as required.

#### Differentiation of human white adipose tissue cells

Human white adipose tissue (hWAT) adipocytes were derived from an established preadipocyte immortalised cell line^104^ generated from human neck fat samples. The wild-type hWAT and single-gene knockdown (including the negative control cell line with the empty Cas9 vector) cells were cultured in Dulbecco’s Modified Eagle Medium (DMEM) containing 10% fetal bovine serum (FBS). To differentiate into adipocytes via addition of differentiation mixture, the cells were induced with 0.5mM 3-isobutyl-1-methylxanthine (IBMX, Sigma Cat #5879) and 0.1μM dexamethasone (Sigma Cat #1756), 33µM Biotin (Sigma Cat # 4639), 17µM Pantothenate (Sigma Cat #5155), 0.5μM insulin (Sigma Cat #2643), 2nM 3,3′,5-Triiodo-L-thyronine (T3, Sigma, Cat #6397), and 1μM Rosiglitazone (APE Cat #A4304) for 3 days. Next, culture medium was replaced to DMEM supplemented with 10% FBS and 33µM Biotin, 17µM Pantothenate, 0.5μM insulin, 2 nM 3,3′,5-Triiodo-L-thyronine, and 1μM Rosiglitazone for 3 days. Medium was subsequently changed every 3 days for the following days. The cells were scrupulously maintained below 70–80% confluence at all assay stages to preserve their differentiation capacity.

#### Lipid staining with BODIPY

Undifferentiated and differentiated hWAT and single-gene knockdown cells were stained with 2.9 μM BODIPY 505/515 (ThermoFisher Cat #4639) and Hoechst (Hoechst 33342, Trihydrochloride, Trihydrate – 10 mg/mL solution in water, ThermoFisher Cat #H3570) for 15 min at 37°C in complete medium, washed with PBS. Cells were analysed for fluorescence intensity using an Opera Phenix™ High-Content Screening System (Revvity, UK) and Harmony® high-content analysis software v4.9 (Revvity, UK) provided by the Cellular High Throughput Screening Group (cmd.ox.ac.uk). For each knockdown condition, there were six replicates of undifferentiated and differentiated cells.

#### RNA sequencing of human white adipose tissue cells

Wild-type hWAT and single-gene knockdown cells were seeded in six-well plates and collected after either (1) zero (undifferentiated), 3, 8 or 24 days of differentiation when assessing sufficient expression in wild-type hWAT or (2) 14 days of differentiation when performing RNA-seq for cell lines involved in the CRISPR knockdown experiment (wild-type hWAT, and single-gene knockdown cells, including Cas9-empty cells). Following collection, RNA was isolated using TRIzol (Thermo Fisher Scientific, Carlsband, CA, USA). Subsequently, RNA samples were purified using the Direct-zol™ RNA Miniprep protocol as per manufacturer’s (Zymo Research, Cat # R2050) recommendations. Quantification of RNA was performed using the Nanodrop 1000 Spectrophotometer. All RNA samples were sequenced using Illumina 2×150bp paired-end sequencing. Read alignment was performed with STAR^105^ v2.7.10b. Transcript quantification estimation was performed with RSEM^106^ v1.3.2. Normalised transcript counts were calculated with DESeq2^70^ v1.42.0.

#### cDNA generation and quantitative RT-PCR

For quantitative RT-PCR (RT-qPCR), total RNA was extracted for gene expression analysis using Direct-zol RNA Miniprep, (Cat #R2050, Zymo Research). After measurement of the RNA (Nanodrop, ThermoScientific, USA), the cDNA synthesis was performed using using High-Capacity cDNA Reverse Transcription Kit (Applied Biosystems™, Cat #4368814) according to the manufacturer’s instructions. RT-qPCR was carried out with 1 μL of the 1:5 diluted reverse transcription sample with 10 μL of 2× SYBR Green Master Mix (iQ SYBR Green Supermix, Cat #1708882, Biorad) and 100 nM specific gene primer pairs in a 10-μL total volume in 96-well microtiter plates. The primer sequences for this study are listed in Supp. Table 11. LightCycler 96 (Roche, USA) was used to measure C_T_ values of each sample. Each experiment was performed at least in triplicate. β-Actin mRNA was employed as an internal standard. The expression level of each gene was determined by RT-qPCR and normalised against β-Actin mRNA level. Knockdown was confirmed by one-sided *t*-test (alpha=0.05) using the 2^-ΔΔCT^ method^107^ for amplification performed in separate wells, with the alternative hypothesis that knockdown decreased gene expression.

#### Differential expression analysis of CRISPR adipocyte RNA sequencing

Differential expression analysis was performed using DESeq2^70^ v1.42.0 on RNA sequencing data from CRISPR adipocytes. Genes with raw count less than 10 were removed. The Cas9-empty cell line was used as the baseline condition. Each knockdown or wild-type hWAT was then separately compared to Cas9 empty.

#### Statistical analysis of lipid accumulation assays

We quantify lipid accumulation using whole-well mean cytoplasm fluorescence of the BODIPY stain, as measured by the Opera Phenix™ High-Content Screening System. For each knockdown and the wild-type hWAT, we combined replicates with the baseline negative control, Cas9 empty, and regressed readout (mean cytoplasm fluorescence) against knockdown status:

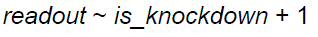

For wild-type hWAT replicates, *is_knockdown* is True to separate Cas9 empty and the wild-type replicates. After regression, we calculated Cook’s distance for each observation. Outliers were removed if Cook’s distance exceeded the median of the *F* distribution with degrees of freedom *n* and *n-p*, where *n* is the number of observations included in the regression (lipid accumulation *n*=12) and *p* is the number of predictors, including the intercept (*p*=2)^108^. Only one observation was flagged as an outlier, for the *SLTM* knockdown: cytoplasm mean fold change relative to Cas9 empty=1.618, Cook’s distance=0.865. We removed this outlier and reran the regression for *SLTM* knockdown lipid accumulation.

We consider the knockdown to have a significant effect on the readout if *P*<0.05 for the *is_knockdown* term from the linear regression. Note that this is the equivalent of a two-sided *t*-test of mean readout between observations from Cas9 empty and a single-gene knockdown or the wild-type hWAT. In Supp. Table 13 we report the fold change of mean fluorescence of each single-gene knockdown relative to the Cas9 empty cell line and the *P*-value from the two-sided *t*-test, after removing outliers identified using Cook’s distance.

#### Druggability evidence

Druggability evidence was collated for each knockdown target gene using OpenTargets profiles^109^, including both known drugs and drug tractability. Here we provide a summary of terms within the “Tractability” columns:

*Small molecule*; Structure with Ligand: Target has been co-crystallised with a small molecule^110^, High-Quality Ligand: Target with ligand(s) (PFI ≤ 7, SMART hits ≤ 2, scaffolds ≥ 2)^111^, High-Quality Pocket: Target has a DrugEBIlity^112^ score of ≥ 0.7, Med-Quality Pocket: Target has a DrugEBIlity^112^ score between 0 and 0.7, Druggable Family: Target is considered druggable as per the Finan *et al.*^113^ druggable genome pipeline.

*Antibody*; UniProt loc high conf: High confidence that the subcellular location of the target is either plasma membrane, extracellular region/matrix, or secretion^114^, GO CC (Gene Ontology Cellular Component) high conf: High confidence that the subcellular location of the target is either plasma membrane, extracellular region/matrix, or secretion^115^, UniProt loc med conf: Medium confidence that the subcellular location of the target is either plasma membrane, extracellular region/matrix, or secretiont^114^, UniProt SigP or TMHMM^116^ (transmembrane helix prediction): Target has a predicted signal peptide or trans-membrane regions, and not destined to organelles (source: Uniprot SigP, TMHMM^116^), GO CC med conf: Medium confidence that the subcellular location of the target is either plasma membrane, extracellular region/matrix, or secretion^115^, Human Protein Atlas loc: High confidence that the target is located in the plasma membrane^117^.

*PROTAC*; Literature: Target mentioned in a set of PROTAC-related publications curated by OpenTargets^109^, UniProt Ubiquitination: Target tagged with the Uniprot keyword “Ubl conjugation [KW-0832]”, which indicates that the protein has a ubiquitination site, based on evidence from the literature^114^, Database Ubiquitination: Target has reported ubiquitination sites in PhosphoSitePlus^118^, mUbiSiDa^119^, or Kim *et al.*^120^, Half-life Data: Target has available half-life data^121^) Small Molecule Binder: Target has a reported small-molecule ligand in ChEMBL with a measured activity of at least 10 μM in a target-based assay^111^.

## Data and code availability

Summary statistics for all phenotypes will be made available through the GWAS Catalog upon publication. All code used in this study will be made available through GitHub upon publication.

## Acknowledgements

We would like to thank Dr. Val Millar and the Ebner group for expert help in the use of PerkinElmer Opera Phenix/High Content Imaging. We also acknowledge wet lab resources from Prof. Benedikt Kessler (Mass Spectrometry for Drug Target Discovery Lab) and Dr. Adan Pinto Fernandez (Translational Ubiquitomics-Cancer Immunology Lab). This research was conducted using the UK Biobank resource under application number 11867. We thank UK Biobank participants for their contribution.

## Research ethics statement

This study used data from the UK Biobank. The North West Multi-centre Research Ethics Committee (MREC) gave approval to UK Biobank as a Research Tissue Bank (RTB) approval. This approval means that researchers do not require separate ethical clearance and can operate under the RTB approval. This study used a human preadipocyte immortalised clonal cell line derived by Xue *et al*.^104^, whose study followed the institutional guidelines of and was approved by the Human Studies Institutional Review Boards of Beth Israel Deaconess Medical Center and Joslin Diabetes Center.

