## Supplementary Notes and Figures for "Combining evidence from human genetic and functional screens to identify pathways altering obesity and fat distribution"

[*,#] shared authorship

### Supplementary notes

#### Note S1. Calculating unweighted burden effect sizes

First, summary statistics for variant-level results are recalculated relative to the minor allele: the sign of the *t*-statistic is flipped if the original effect allele is not the minor allele and the frequency of the effect allele is recalculated accordingly. Then, for all single-variant and grouped ultra-rare variant burden results in a given gene which satisfy the maximum minor allele frequency and variant consequence of the desired result to unweight, the following summary statistics are summed: *t*-statistic, variance of *t*-statistic, and minor allele frequency. The unweighted *t*-statistic is calculated by dividing the summed *t*-statistics by the summed variance of *t*-statistics. The unweighted minor allele frequency is the summed minor allele frequency.

$$t_{unweighted,\,gene} = \frac{\sum_j t_{weighted,\,j}}{\sum_j Var(t_{weighted,\,j})}$$

$$MAF_{unweighted,\,gene} = \sum_j MAF_{weighted,\,j}$$

#### Note S2. Significant genes with low minor allele count

In sex-combined and sex differential gene-level associations we sought to avoid spurious associations by excluding putatively significant genes if they were supported by a total minor allele count (MAC) <10 across both sexes.

Among the sex-combined gene-level associations which were significant at FDR≤1% (SKAT-O $P \leq 4.37\times10^{-5}$), there were seven genes which were excluded for having MAC<5: *DEFB112* (MAC=1), *CHMP4B* (MAC=2), *FEZF2* (MAC=3), *GLP1R* (MAC=3), *PCBD2* (MAC=4), *VGF* (MAC=8), and *TM4SF20* (MAC=9) (Supp. Table 3).

In the sex differential analysis, we excluded a significant (sex-difference $P<2.67\times10^{-6}$) differential effect at *SENP5* for visceral adipose tissue volume (female beta=0.08930 (0.028055), male beta=-0.09967 (0.028675), sex-difference $P=2.33\times10^{-6}$) due to combined MAC<10 (female MAC=2, male MAC=2) (Supp. Table 5).

### Supplementary tables

**Table S1. Sample removal based on sample outliers defined by MAD thresholds** (median ± 4 MADs), split by sequencing tranche.

| QC Metric | # Samples in UK Biobank Whole Exome Sequencing Tranche |
|---|---|

| | 250k | 150k | 50k |
|---|---|---|---|
| **Total before QC** | **237,091** | **139,745** | **46,034** |
| Number of deletions | 1,510 | 825 | 344 |
| Number of insertions | 1573 | 1,118 | 236 |
| Number of SNPs | 1,856 | 1,013 | 362 |
| Ratio of insertions to deletions | 1,741 | 954 | 339 |
| Ratio of transitions to transversions | 2,890 | 1,598 | 1781 |
| Ratio of heterozygous variants to homozygous alternate variants | 2,301 | 1,496 | 478 |
| Total failing | 10,861 | 6,396 | 3,238 |
| **Total passed QC** | **226,230** | **133,349** | **42,796** |
| **Grand total** | | | **402,375** |

**Table S2. Sample sizes and phenotype standard deviations for each adiposity-related trait (sex-combined and sex-stratified)**

| Phenotype | UKB Field ID | Both sexes | | Female | | Male | |
|---|---|---|---|---|---|---|---|
| | | N | Std. dev. | N | Std. dev. | N | Std. dev. |
| Body mass index (BMI) | 21001 | 401059 | 4.74783 | 217716 | 5.11381 | 183343 | 4.22661 |
| Waist-to-hip ratio, adjusted for BMI (WHRadjBMI) | n/a | 400948 | 0.08064 | 217655 | 0.06263 | 183293 | 0.05214 |
| Body fat percentage (BF%) | 23099 | 395147 | 8.50612 | 214846 | 6.86752 | 180301 | 5.80123 |
| Android tissue fat percentage | 23247 | 39671 | 0.11148 | 20456 | 0.11721 | 19215 | 0.10322 |
| Gynoid tissue fat percentage | 23264 | 39671 | 0.09288 | 20456 | 0.06664 | 19215 | 0.06159 |
| Android-gynoid tissue fat percentage ratio | n/a | 39671 | 0.08305 | 20456 | 0.07372 | 19215 | 0.06631 |
| Total tissue fat percentage | 23281 | 39671 | 0.28970 | 20456 | 0.20838 | 19215 | 0.24515 |
| Visceral adipose tissue (VAT) volume | 22407 | 21253 | 2.28788 | 11000 | 1.52205 | 10253 | 2.35928 |
| Abdominal fat ratio | 22434 | 20687 | 0.11222 | 10892 | 0.10543 | 9795 | 0.09907 |

For Supplementary Tables 3-16, refer to Excel file.

**Table S3. Gene-level results for all FDR≤1% significant genes**
**Table S4. Replication of UKB-significant genes in All of Us**
**Table S5. Phenome-wide associations from Genebass**

**Table S6. Sex-specific and sex-differential gene-level results.** These are genes that only reach significance in sex-specific strata and genes with significant heterogeneity in effect sizes between sexes (phenotype, male-specific beta & SE & P, female-specific beta & SE & P, sex-heterogeneity P-value)

**Table S7. Obesity age-of-onset longitudinal analysis using Cox proportional hazards model**

**Table S8. Combined evidence table (hWAT counts, literature review)**

**Table S9. Guide RNAs per gene target**

**Table S10. RNA-seq differential expression to confirm knockdown**

**Table S11. PCR primers**

**Table S12. RT-qPCR differential expression to confirm knockdown**

**Table S13. Effect of knockdown on lipid accumulation**

**Table S14. Evidence of druggability for knockdown target genes**

**Table S15. Genes differentially expressed between each knockdown and Cas9-empty cell line, with logFC and P-value**

**Table S16. GSEA pathway analysis for RNA-seq of knockdowns**

### Supplementary figures

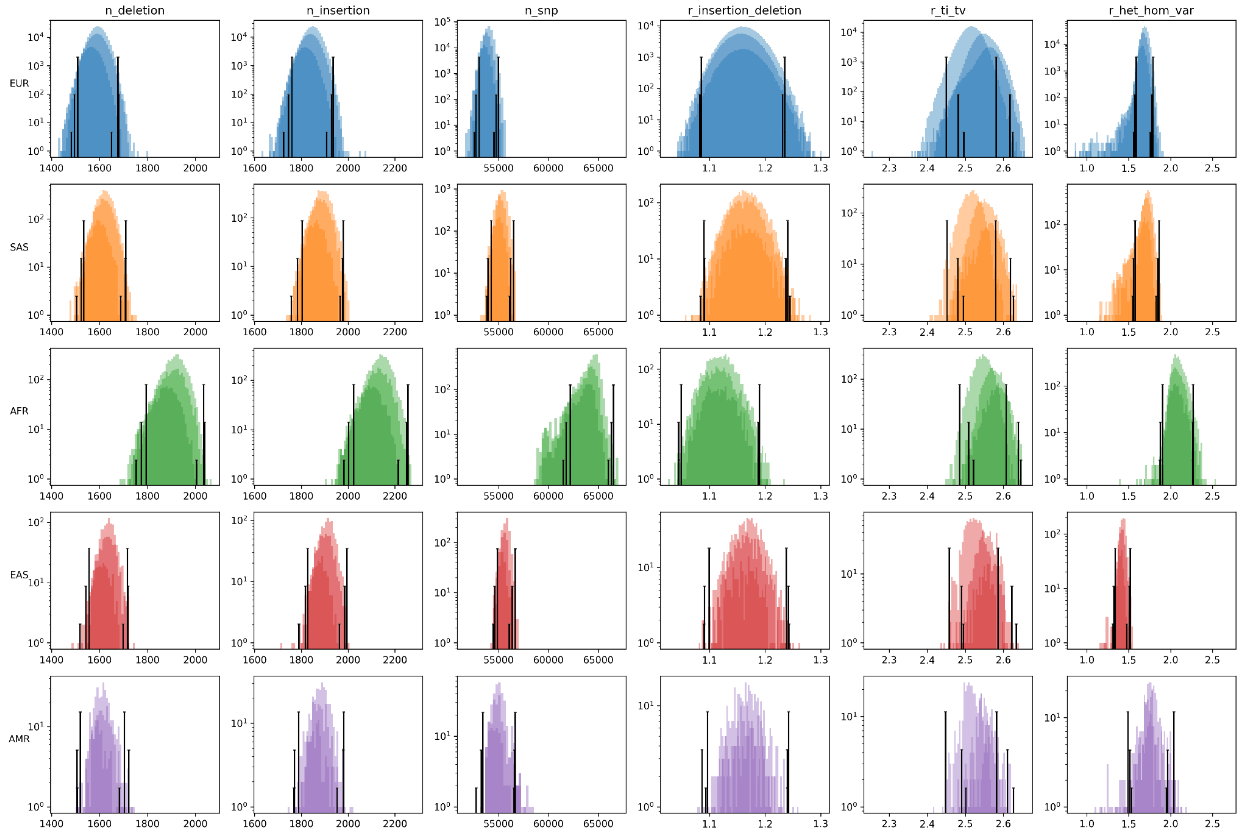


**Figure S1. MAD thresholds, split by tranche**. Samples with any of n_deletion, n_insertion, n_snp, r_insertion_deletion, r_ti_tv, and r_het_hom_var exceeding four MADs from the median are removed. MAD thresholds are displayed as vertical lines, conditional on tranche size (50k, 200k, 250k).

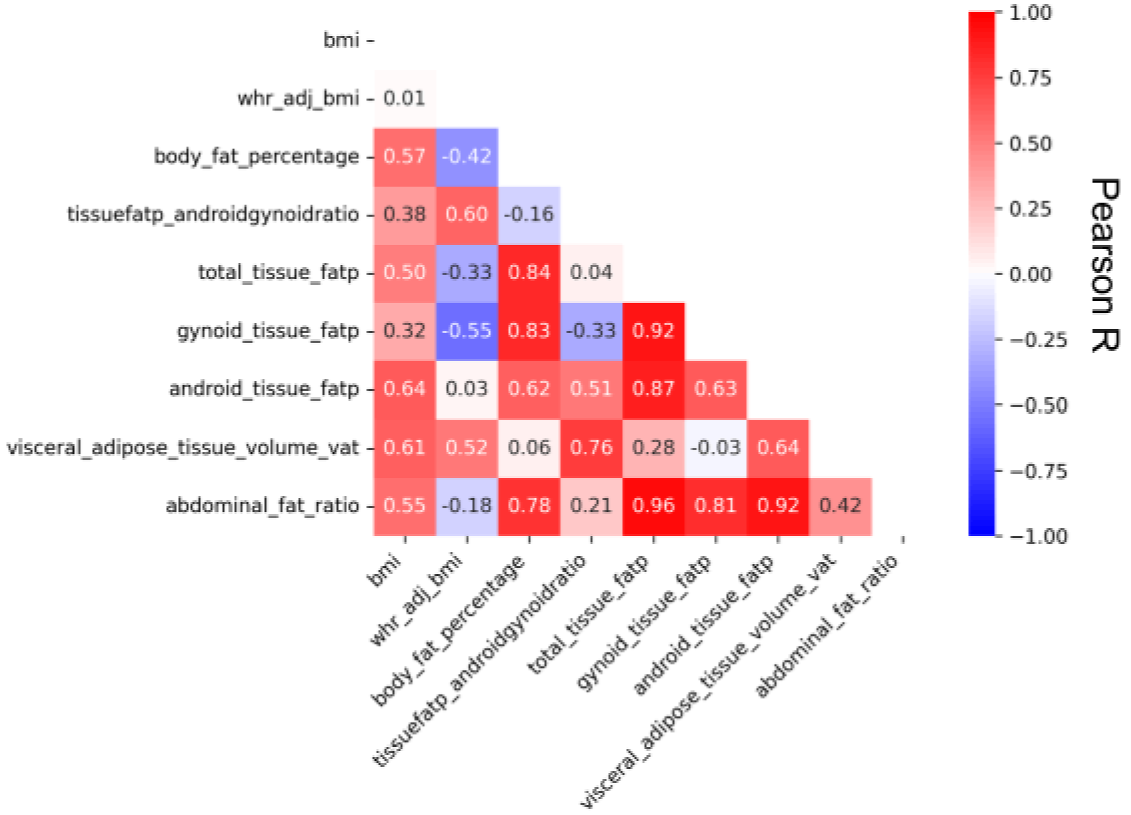


**Figure S2. Obesity and fat distribtuion trait correlation heatmap**. Correlations for this heatmap were calculated using the subset of individuals used for our genetic association testing. By definition of WHRadjBMI, correlation between BMI and WHRadjBMI should be zero, but here it is nonzero (R=0.01) because WHRadjBMI was calculated on a superset of individuals used for the correlations.

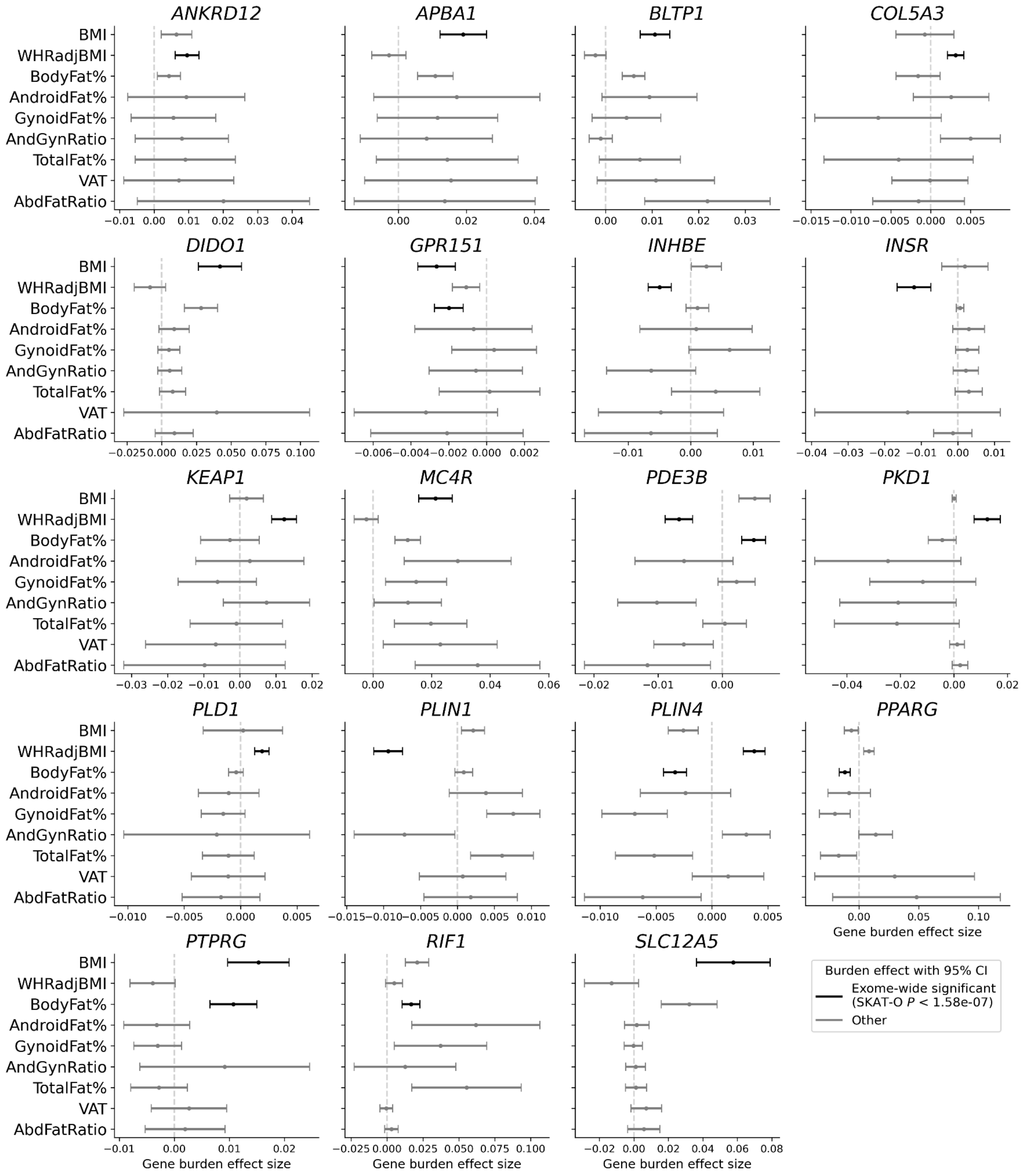


**Figure S3. Gene burden effects across all nine obesity and fat distribution traits for 19 genes with exome-wide significant burden associations.** Confidence intervals for effect size defined as +/-1.96 standard errors. Only the result of the consequence mask with the lowest SKAT-O *P* is shown for each trait-gene pair. The vertical dotted line marks an effect size of zero. VAT, visceral adipose tissue.

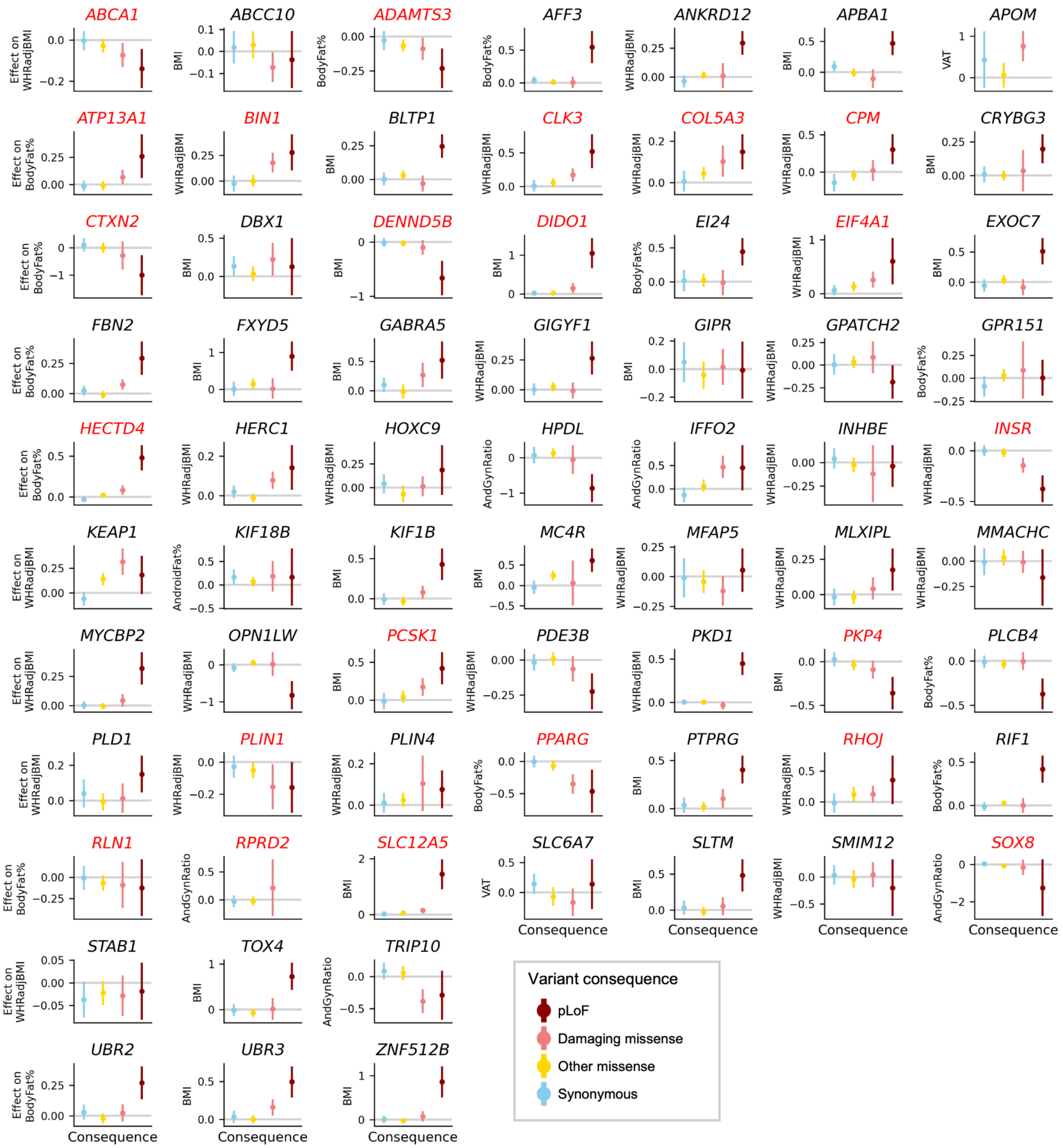


**Figure S4. Ultra-rare variant allelic series for all FDR≤1% significant genes.** Monotonic allelic series are highlighted with red gene symbols. Allelic series are defined as monotonic if they have strictly increasing or strictly decreasing effect sizes across increasingly deleterious variant consequences (least to most deleterious: synonymous, other missense, damaging missense, pLoF). Effect sizes come from ultra-rare variant burden for variants with minor allele count≤10 in each gene, grouped by variant consequence. Confidence intervals for effect size defined as +/-1.96 standard errors. pLoF, predicted loss-of-function.

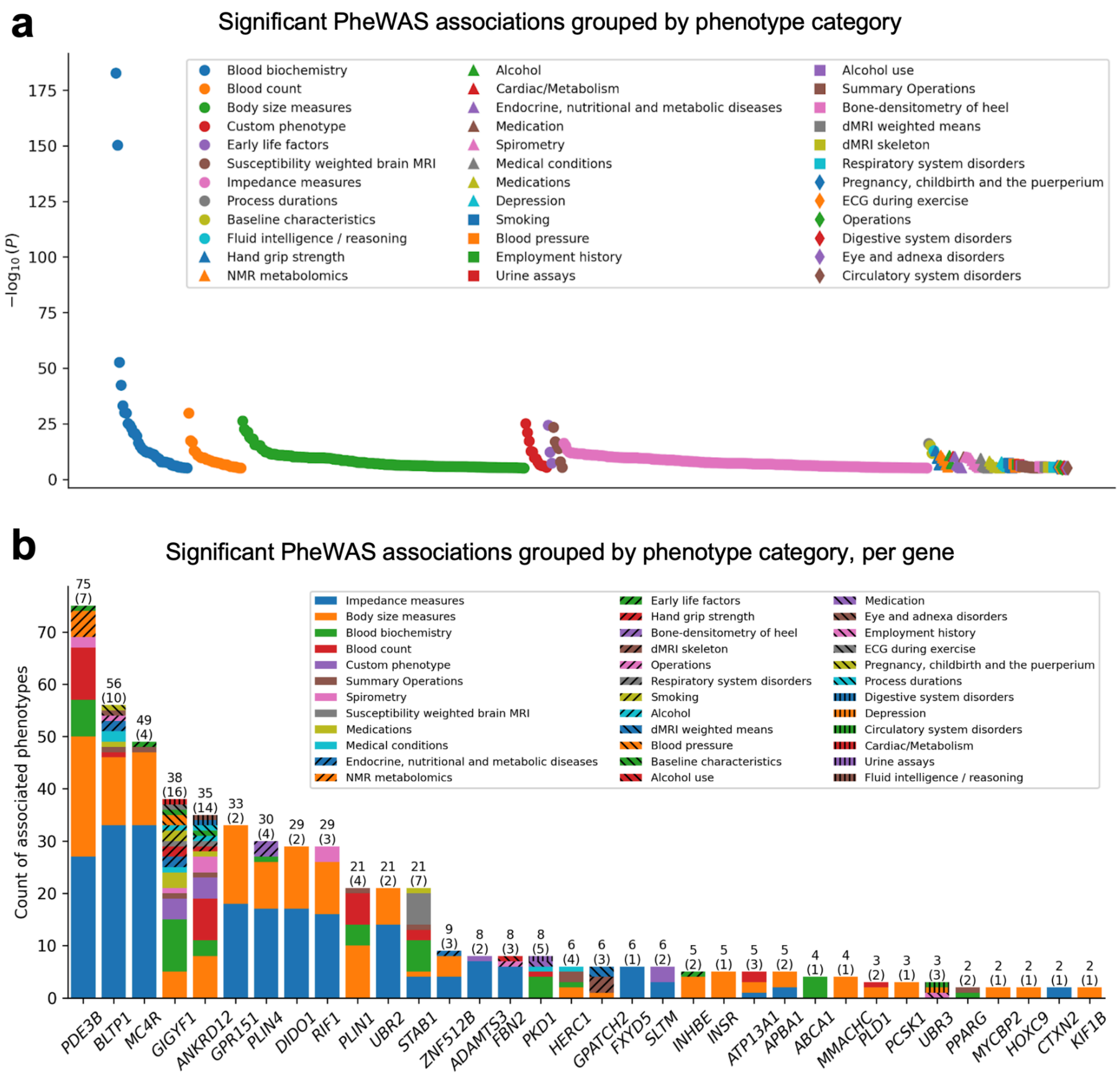


**Figure S5. PheWAS of obesity and fat distribution associated genes using Genebass summary statistics.** Only the pLoF variant mask results from Genebass are used. Significant associations are controlled for FDR≤1% (SKAT-O $P \leq 9.98 \times 10^{-6}$), resulting in 549 significant associations across 211 phenotypes and 41/69 obesity and fat distribution associated genes. **a,** Significant associations grouped by phenotype category, with phenotype groups ordered from left to right by the lowest *P*-value in the category. Significance of association is measured on the y-axis as $-\log_{10}$(Genebass SKAT-O *P*-value). **b,** Significant associations per gene, grouped by phenotype category. The total number of significant phenotype associations is shown at the top of each bar, with the number of phenotype categories shown in parentheses. Only genes with at least two phenotype associations are shown.

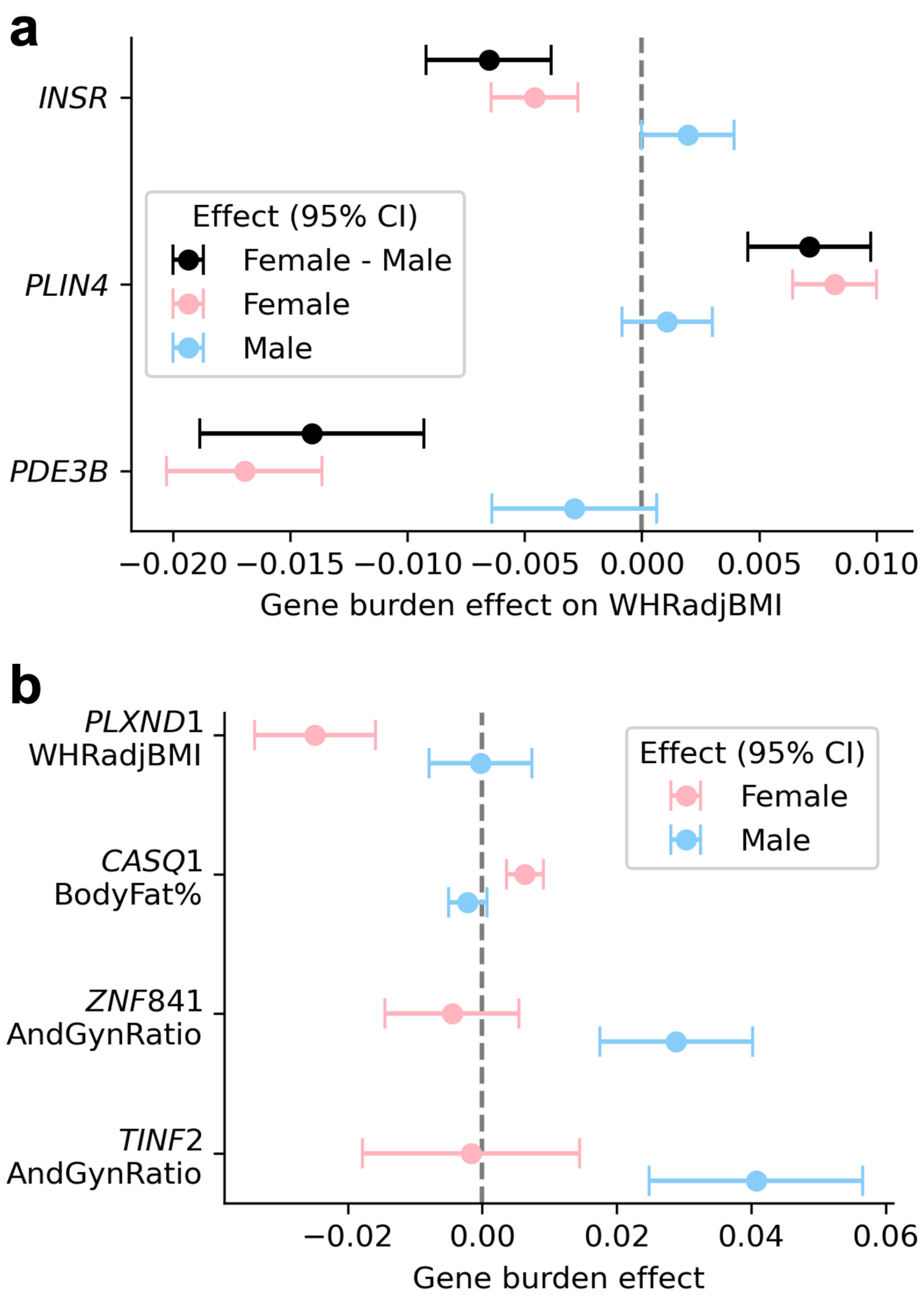


**Figure S6. Sex-differential and sex-specific analysis. a,** Genes with significant sex-differential effects (sex-difference $P<2.67\times10^{-6}$, Bonferroni adjusted for 18,737 genes tested for sex-differential effects). All three significant sex-differential gene burden effects are on WHRadjBMI. **b,** Female- (*PLXND1, CASQ1*) and male-specific (*ZNF841, TINF2*) gene-level significant associations ($P<2.67\times10^{-6}$, Bonferroni adjusted for 18,737 genes tested for sex-specific effects). Confidence intervals for effect size defined as +/-1.96 standard errors.

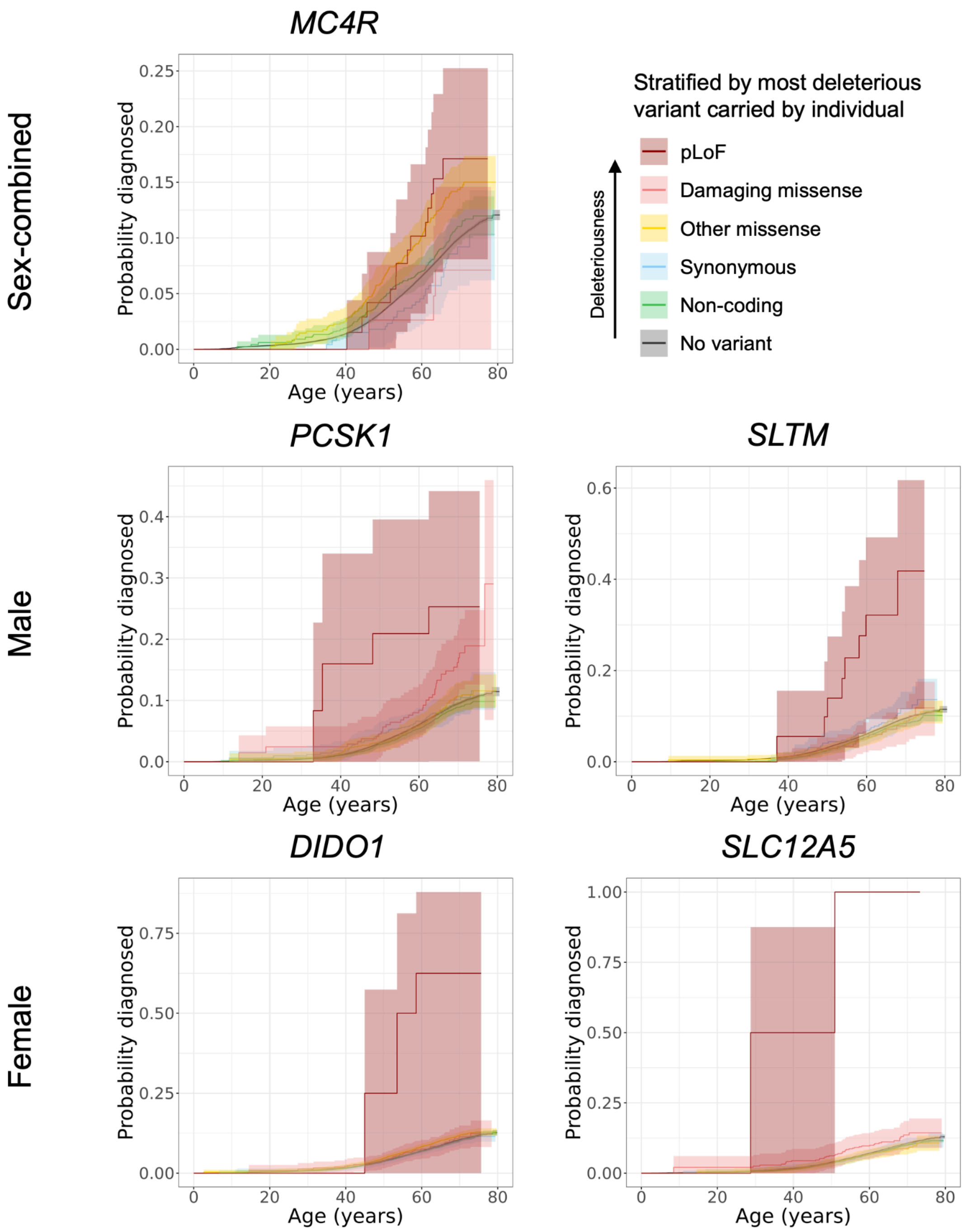


**Figure S7. Longitudinal obesity age-at-onset analysis.** Longitudinal analysis was performed using Cox-proportional hazards modelling (Methods). Individuals are stratified by the most deleterious carried in the gene. Only genes with significant age-of-onset associations in both sexes (*MC4R*), males only (*PCSK1, SLTM*), or females only (*DIDO1, SLC12A5*) are shown. 95% confidence intervals are indicated by shading.

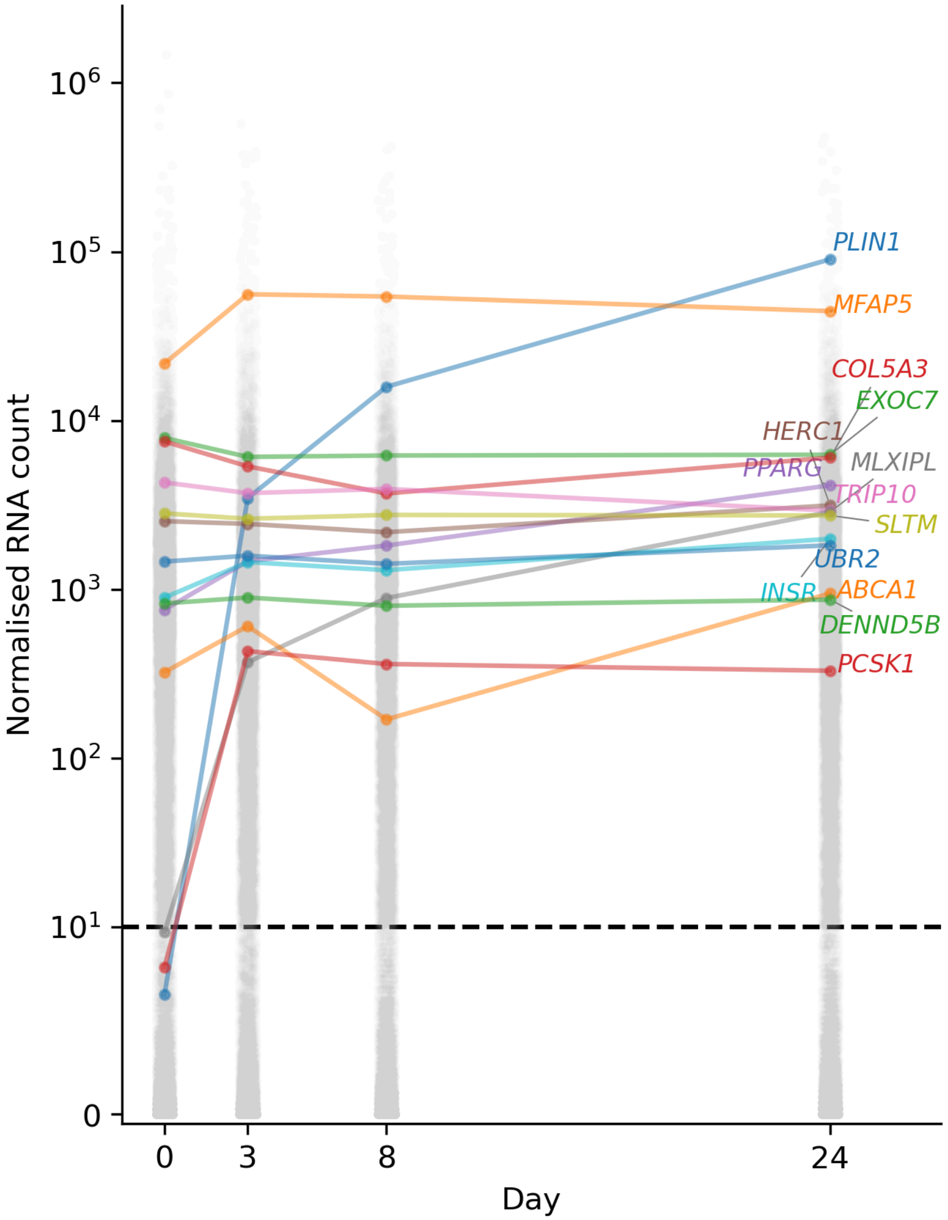


**Figure S8. Longitudinal mRNA expression in wild type human white adipose tissue of genes selected for knockdown.** Normalised mRNA count was measured at four time points across 24 days of differentiation. Day 0 corresponds to the undifferentiated state. Colored traces are genes selected for knockdown. All other genes are indicated with grey points. The horizontal dashed line indicates the minimum threshold needed on days 8 and 24 for a gene to be selected as a knockdown target. The y-axis uses the ‘symlog’ scale, such that the scale is linear between 0 and 10 and logarithmic for values greater than 10.

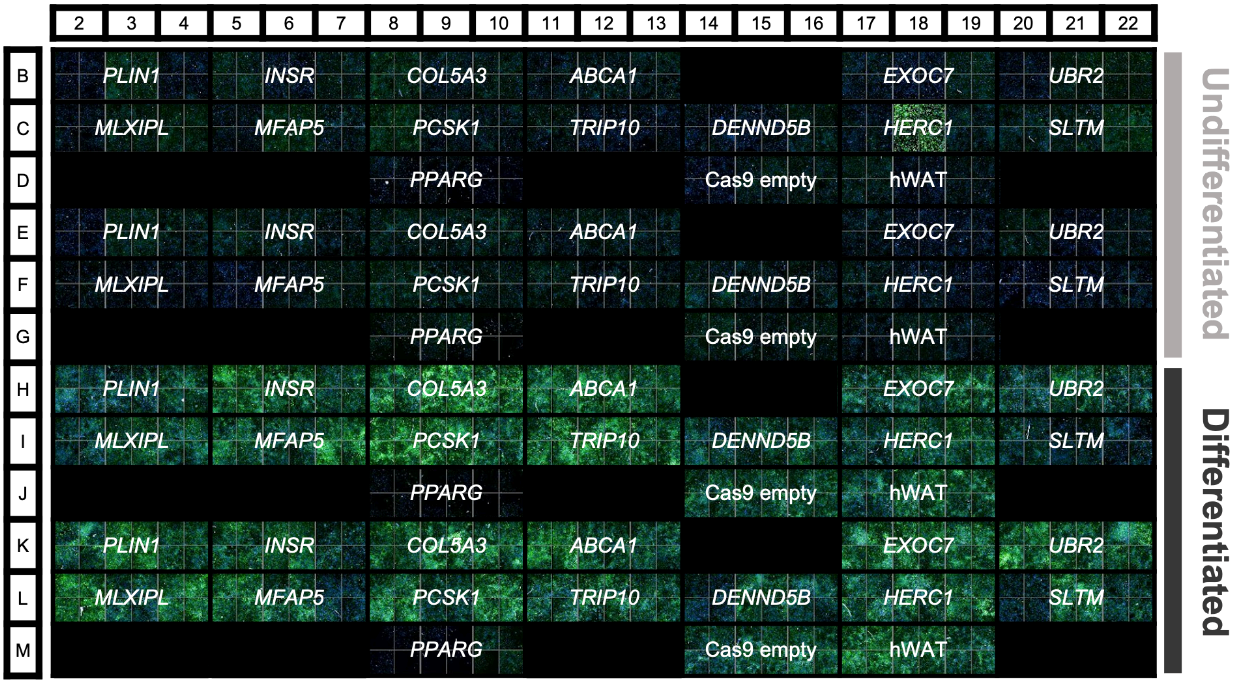


**Figure S9. Imaging of well plates for lipid accumulation knockdown adipocytes.** Lipid accumulation is highlighted in green by fluorescent BODIPY staining. Each knockdown cell line had six replicates for each differentiation status (undifferentiated, differentiated).
